# Pathogenic Mitofusin 2 variants causing myopathy drive endosomal mtDNA release and inflammation

**DOI:** 10.64898/2026.09.22.26363493

**Authors:** Mashiat Zaman, Joshua David, Ayshin Mehrabi, Cole Chute, Jingti Deng, Tyler Soule, R. Luke Wiseman, Costin N. Antonescu, Pina Colarusso, Gerald Pfeffer, Timothy E. Shutt

## Abstract

Pathogenic variants in Mitofusin 2 (MFN2) cause the peripheral neuropathy Charcot-Marie-Tooth Type IIA (CMT2A), with a subset of patients presenting with myopathy. Yet how MFN2 dysfunction causes these phenotypes remains mechanistically undefined. Here, we identify a conserved pathomechanism in MFN2-linked myopathy variants, where mitochondrial DNA is released into RAB5 early endosomes, robustly activating TLR9 and cGAS-STING inflammatory signaling. We further show that MFN2 dysfunction drives two mechanistically distinct pools of extra-mitochondrial mtDNA, one in endosomes and the other present mitochondrial-derived vesicles (MDVs). These two pools of released mtDNA can be distinguished based on their size, with endosomal mtDNA forming larger puncta than mtDNA present in MDVs. Finally, we show that pharmacological activation of the integrated stress response reduces both pools of extra-mitochondrial mtDNA. Together, our work defines a conserved cellular phenotype linking MFN2 dysfunction to myopathy and suggests a tractable therapeutic strategy for mtDNA release-associated pathologies.

**Graphical Abstract:** 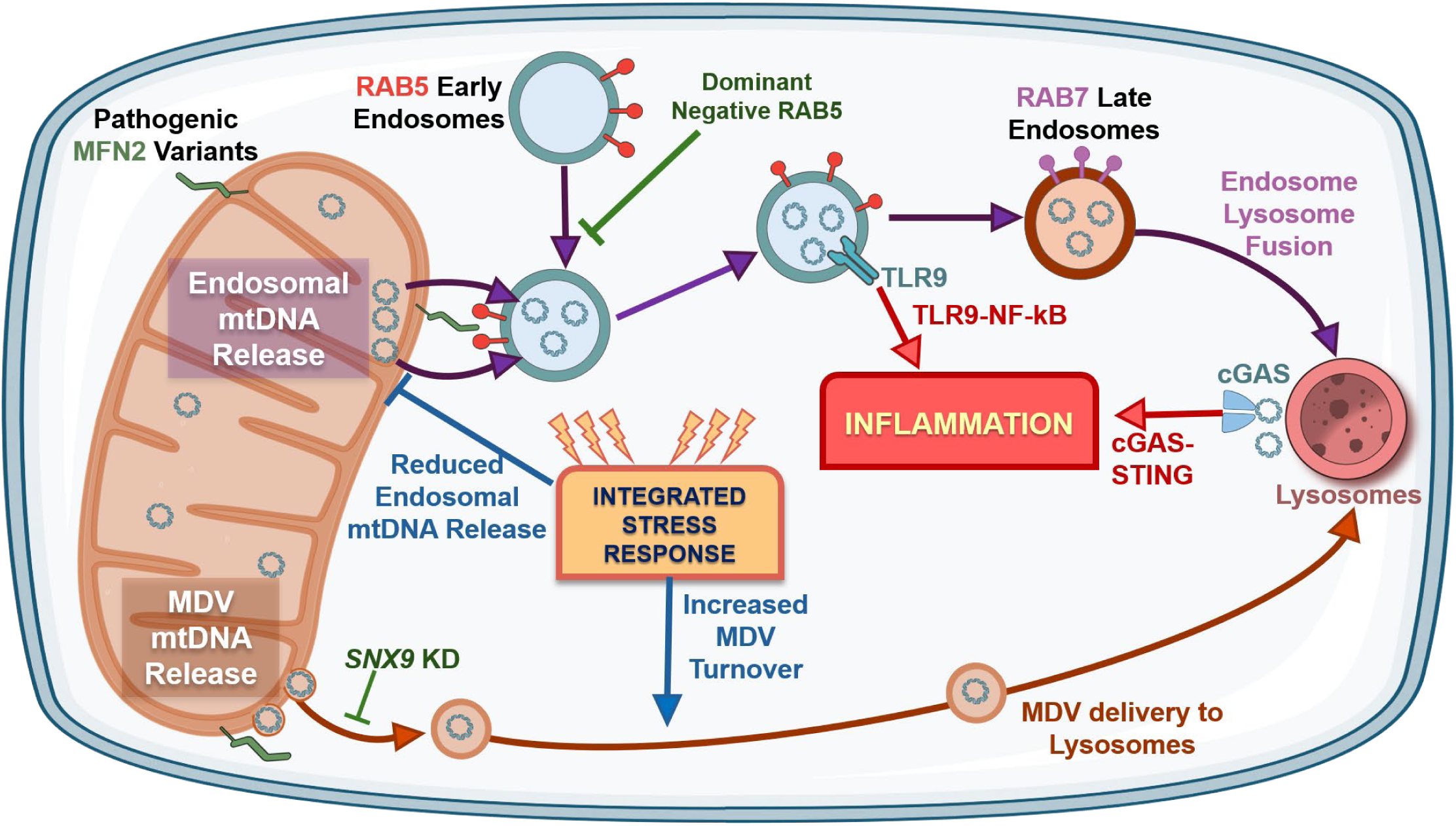

## Introduction

Mitofusin 2 (MFN2) is an integral outer-mitochondrial-membrane (OMM) GTPase, discovered over two decades ago for its role in regulating mitochondrial fusion (Chen et al., 2003; Hales & Fuller, 1997). Since then, we have come to appreciate that MFN2 has several other functions, as it is also involved in mitochondrial organelle contact sites, mitochondrial autophagy, mitochondrial motility and regulation of the mitochondrial genome (Zaman & Shutt, 2022). Pathogenic MFN2 variants cause disease, predominantly the hereditary axonal peripheral neuropathy Charcot-Marie-Tooth disease 2a, with over 100 pathogenic variants in MFN2 linked to CMT2a (Pipis et al., 2020). However, a subset of MFN2 variants are linked to pathologies beyond neuropathy, including myopathies (Brožková et al., 2013; Choi et al., 2015; Chung, 2006; Estilow et al., 2012; Franco et al., 2023; Hayashi et al., 2023; Hines et al., 2023; Irazoki et al., 2023; Rouzier et al., 2012; Sawyer et al., 2015; Zaman et al., 2025). Nonetheless, the mechanisms by which MFN2 dysfunction cause this clinical heterogeneity remains poorly understood.

Although it was once thought that myopathy in CMT2a patients was downstream of peripheral neuropathy, recent work shows that muscle pathology due to MFN2 dysfunction can manifest without peripheral neuropathy. For example, the MFN2 L643P variant was identified in a mouse model exhibiting myopathy without peripheral neuropathy (Hines et al., 2023). Meanwhile, our recent work described the novel Q367H MFN2 variant in a late-onset patient with myopathy independent of the neuropathy (Zaman et al., 2025). While these examples show that myopathy and neuropathy phenotypes can be independent, the underlying mechanisms causing these phenotypes remain uncertain. Focusing on myopathy, it is notable that muscle-specific MFN1 knockout causes myopathy mediated by mtDNA TLR9 activation (Irazoki et al., 2023). While previous work also connected the Q367H MFN2 variant to mtDNA release and TLR9 and cGAS-STING inflammation (Zaman et al., 2025), it remains unknown whether other MFN2 variants also activate mtDNA-mediated inflammation, or if this inflammation contributes to pathology.

While functional studies show that MFN2 depletion leads to mtDNA release and subsequent inflammation (Irazoki et al., 2023), how mtDNA is release occurs is not fully understood for MFN2 dysfunction. General mechanisms of mtDNA release can be divided into pore-mediated or vesicle-mediated processes (Newman & Shadel, 2023; VanPortfliet et al., 2024). Pore-mediated mechanisms include BAX/BAK-dependent release during apoptosis (McArthur et al., 2018) and mPTP/VDAC-mediated release during oxidative stress (Xian et al., 2022), and release of mtDNA directly into the cytosol, where it activates cGAS-STING signaling (Lepelley et al., 2021). Meanwhile, vesicular pathways of mtDNA release include SNX9-mediated mitochondrial-derived vesicles (MDVs) (VanPortfliet et al., 2024) and RAB5-early endosomal compartments that are recruited to mitochondria (Newman et al., 2024). Notably, both vesicular release pathways can ultimately deliver mtDNA to lysosomes, where it can subsequently be released into the cytosol via gasdermin pores (Nguyen et al., 2025) and detected by cGAS-STING. However, when mtDNA is released via endosomes, it is also detected by the Toll-like Receptor 9 (TLR9) signalling pathway (Fang et al., 2016; Irazoki et al., 2023; Zaman et al., 2025), which would otherwise be bypassed by MDV release. Therefore, the mechanism of the mtDNA release plays a fundamental role in activating distinct innate immune signaling pathways. In this regard, how MFN2 dysfunction affects the different mechanisms of mtDNA release and the subsequent downstream inflammation remains unknown.

To better understand the clinical relevance of mtDNA-mediated inflammation in MFN2 disease, we screened a panel of ten pathological MFN2 variants linked to a variety of pathologies, including myopathy. Notably, we observed that mtDNA release is associated with all MFN2 variants that we investigated, with a clear correlation between variants linked to myopathy and mtDNA release into early endosomes. Furthermore, we demonstrate that MFN2 dysfunction can also promote mtDNA release via MDVs. As such, we describe two distinct mechanisms of mtDNA release and downstream inflammation that can occur in response to MFN2 dysfunction. Finally, we outline how pharmacological activation of the integrated stress response reduces mtDNA outside the mitochondrial network, providing a possible therapeutic approach for MFN2-mediated myopathy.

## Materials and Methods

### Cell Lines and Cell Culture

MFN2 KO cells were a generous gift from Dr. Edward A Fon (McGill University, Canada). U2OS cells were maintained with DMEM media (Thermo Fisher Scientific, 11965118) supplemented with 10% heat-inactivated FBS (Thermo Scientific, A5256701) and incubated at 37°C and 5% CO_2_. The KO re-expression cell lines were generated by viral transduction, using viruses made from Phoenix Cells (ATCC, CRL3213) and the plasmids were generated using VectorBuilder, to carry point mutations of interest in the MFN2 open reading frame. The Phoenix cells were transfected using Lipofectamine 3000 (Thermo Fisher Scientific, L3000015) and virus was harvested for three consecutive days, concentrated (Takara Bio, 631232), then added to the MFN2 KO cells. Transduced cells were sorted for target fluorescence using a SONY SH800 cell sorter at the University of Calgary Flow Cytometry Facility. Re-expression was confirmed by western blots to ensure comparable levels of expression across variants and compared to U2OS WT cells. The Conjoint Health Research Ethics Board of the University of Calgary gave ethical approval for the use of patient fibroblasts in this work. Fibroblasts were maintained in DMEM media supplemented with 10% FBS and incubated at 37°C and 5% CO_2_.

### Western Blot Analysis and Antibodies for WB

Western blot analyses were performed as previously reported (Zaman et al., 2025). Briefly, cell pellets were harvested with trypsin (VWR, CA45000-664) from 10cm dishes and lysed using RIPA buffer (Fisher Scientific, 89901) containing 1% protease inhibitor (VWR, 97063-010). Protein concentrations were quantified using a BCA assay with BSA standards (Fisher Scientific, PI23208) and 30µg of protein was loaded into each well of a 10% SDS-PAGE gel. The proteins were run on the gel and subsequently transferred to a PVDF membrane (Bio-Rad, 1620177). The membranes with protein were then blocked with 5% milk in TBS-T for 1 hour at room temperature and primary antibodies were added overnight. Antibodies used were rabbit anti-MFN2 (Thermo Scientific, 711803), anti-alpha-Tubulin (DSHB, 12G10), anti-SNX9 (abcam, ab181856), anti-Rabbit HRP (NEB, 7074S) and anti-Mouse HRP (Thermo Scientific, 31430). Secondary antibodies were incubated for one hour and the membrane was subsequently imaged with ECL (Fisher Scientific, PI34095), using a BioRad ChemiDoc imaging system.

### Immunostaining and Imaging

Immunostaining was performed as previously described (Uddin et al., 2026). Briefly, 20,000 cells were seeded on coverslips in a 24-well plate and allowed to grow for 24 hours. The cells on the coverslips were then washed three times with 1x PBS and fixed with 4% PFA (Electron Microscopy Sciences, 15710) for 20 minutes at 37°C. Cells were then washed three times with 1X PBS and permeabilized with 0.25% Triton-X in 1X PBS for 20 minutes and blocked in 10% FBS in 1X PBS for 1 hour. Primary antibodies were then added in the blocking/permeabilizing buffer and incubated overnight. Antibodies used were: rabbit anti-TOMM20 (Abcam, ab186735), mouse anti-TOMM20 (Santa Cruz Biotechnology, sc-17764), chicken anti-TOMM20 (AVES labs, TOMM20-0100), anti dsDNA (DSHB, AB10805293), anti RAB5 (Cell Signaling Technology, 3547S), anti-RAB7 (Cell Signaling Technology, 9367S), anti-PDH (abcam, AB110333), anti-HA (AVES labs, ET-HA100), anti-mouse 405 (Invitrogen, A482B7), anti-mouse 488 (Thermo Fisher Scientific, A11029), anti-mouse 568 (Thermo Fisher Scientific, A11004), anti-mouse 647 (Thermo Fisher Scientific, A31571), anti-rabbit 488 (Thermo Fisher Scientific, A11034), anti-rabbit 568 (Thermo Fisher Scientific, A11011), anti-rabbit 647 (Thermo Fisher Scientific, A21245), anti-chicken 488 (Thermo Fisher Scientific, A11039) and anti-chicken 568 (Thermo Fisher Scientific, A11040).

Following overnight incubation, secondary antibodies were added for 1 hour and cells were subsequently mounted on glass slides using ProLong Glass Antifade Mountant (Invitrogen, P36965). Fixed cell imaging was acquired using the Olympus Spinning Disk Confocal System (Olympus SD-OSR) was used with a 60× oil immersion objective. For live cell imaging, 35,000 cells were seeded on live cell imaging dishes (Cellvis, D35-20-1.5-N) and grown for 24 hours. Cells were then imaged on the same microscope but maintained at 37OC and 5% CO2 using a CellVivo live cell imaging system.

### Live-cell imaging of mtDNA release dynamics and mitochondria–endosome contacts

For live-cell imaging of mtDNA release dynamics, cells stably expressing mt-HI-NESS (David et al., 2026) were stained with MitoView Green (Biotium-#70054) for 20–25 min under standard culture conditions, washed twice with PBS, and maintained in regular growth medium for imaging. Cells were imaged on a spinning-disk confocal microscope equipped with temperature and CO environmental control. Prior to acquisition, cells were equilibrated in the imaging chamber for approximately 30 min to minimize drift.

Images were acquired using a 60×/1.5 NA objective. Z-stacks were collected around the central plane of the cell, extending several microns above and below the selected focal plane. Z-drift compensation/autofocus was used during acquisition to maintain focal stability. Time-lapse images were acquired at 10 s or 30 s intervals, or as indicated in the figure legends.

For mitochondria–endosome contact imaging, cells were transiently transfected with Emerald-RAB5a (Addgene plasmid #54243). Forty-eight hours after transfection, mitochondria were labeled with MitoTracker Deep Red for 20–25 min under standard culture conditions, washed twice with PBS, and returned to regular growth medium. Live-cell imaging was performed using the same microscope configuration and acquisition settings described above.

### Quantification of mtDNA release dynamics from live-cell imaging

To quantify mtDNA release dynamics from live-cell time-lapse datasets, mt-HI-NESS-positive mtDNA structures were segmented and tracked in Fiji using the TrackMate (Ershov et al., 2022; Tinevez et al., 2017) plugin . Briefly, image sequences were background-corrected and projected over time to enable frame-by-frame detection of mt-HI-NESS-positive puncta. Consistent segmentation and tracking parameters were applied across comparable datasets. Mitochondrial fluorescence was used to generate a mitochondrial mask, which was then used to determine whether tracked mt-HI-NESS-positive objects were mitochondria-associated or extra-mitochondrial.

A release event was conservatively defined as the first frame in which a newly detected mt-HI-NESS-positive object appeared outside the mitochondrial mask and persisted for a minimum track duration. Objects present in preceding frames, objects located at the image boundary, and objects already assigned to a previous release event were excluded to minimize false-positive event detection. For each qualifying release event, the release frame, object birth and death frames, time to release, post-release persistence, and total track lifetime were recorded. Release-event counts were then summarized over time to quantify mtDNA release dynamics.

### Knockdown of SNX9 and Inactivation of RAB5

The knockdown of *SNX9* was performed using TriFECTa DsiRNA kit (IDT, hs.Ri.SNX9.13), transfected into cells using lipofectamine RNAi MAX (Fisher Scientific, 13778075). Knockdowns were verified using Western blot analyses. The mCherry Rab5 (Addgene, 55126) and dominant negative Rab5 (Addgene, 35139) were introduced into the cell by transfection using Lipofectamine 3000.

### Drug Administration

Pharmacological activation of the ISR was performed as previously described (Baron et al., 2025; Bora et al., 2025). Briefly, Halofuginone was added at 100nM (Sigma, 50-576-3001), Parogrelil was added at 10µM (Axon Medchem, 139145-27-0) and ISRIB was added at 200nM (Sigma, SML0843) were reconstituted in DMSO and added to the cells on live cell imaging dishes, before imaging after 6 hours. Bafilomycin was added at 100nM (Abcam, AB120497) in the ISR experiments was added two hours before adding the ISR activators. The compounds used to block the pore-mediated mtDNA release pathways-BAI1 (Cedarlane Labs, HY-103269) and Cyclosporin A (Cedarlane Labs, HY-B0579) were added to live cells for 24 hours before imaging the cells.

### Inflammation and mtDNA Copy Number

qPCR for mtDNA copy number was performed as previously described (Al Khatib et al., 2022). Briefly, total DNA was isolated using a PureLink Genomic DNA Mini Kit (Thermo Fisher Scientific, K182001), as per the manufacturer’s instructions. 100ng of genomic DNA was then analyzed by qPCR using SYBR Green (Thermo Fisher Scientific, A25742) and the delta delta Ct method. The readout for inflammation was performed as previously described (Zaman et al., 2025). Briefly, total RNA was extracted using the HP RNA isolation kit (VWR, CA101414-852) and converted to cDNA using the Advanced cDNA Synthesis Kit (Bio-Rad,1725038). qPCR was performed with 50ng cDNA, using the QuantStudio 6 Real-Time PCR system (Thermo Fisher Scientific). Primer sequences used have previously been reported (Zaman et al., 2025).

### Image Analysis

Confocal microscopy images were imported into FIJI (Schindelin et al., 2012). All images were z-projected before analysis. Release of mtDNA puncta was quantified by repurposing the ‘MitoQC’ plugin on FIJI (Montava-Garriga et al., 2020), treating the released mtDNA as ‘mitolysosomes’ in the plugin. The number of mtDNA in early endosomes was also counted by masking the early endosomes, then overlaying the mtDNA puncta onto that mask and running ‘analyse particles’ on FIJI. The number and size of mtDNA nucleoids, as well as endosomes were also analyzed through the same plugin. MDVs containing mtDNA and PDH were counted manually, but the MitoQC plugin was used to quantify PDH-only or TOMM20-only MDVs. Mito-Endosome distances were calculated using the ‘Nearest Neighbour Distance’ plugin on FIJI. Mitochondrial network morphology was quantified as previously described (Zaman et al., 2026), using the MiNA plugin on FIJI (Valente et al., 2017).

### Statistics, results and image icons

All graphs were made, and statistical analysis was performed in GraphPad Prism. Icons used in the graphical abstract were made using open-source icons from NIH BioArt and Servier Medical Art.

## Results

### Multiple pathogenic MFN2 variants initiate mtDNA release

While MFN2 depletion leads to mtDNA release and inflammation (Irazoki et al., 2023; Zaman et al., 2025), to date only a single pathogenic MFN2 variant, Q367H, has been investigated for this phenomenon (Zaman et al., 2025). Two key challenges for understanding MFN2 pathomechanisms are the lack of patient cells from these rare disease patients, as well as the different genetic backgrounds of MFN2 patients, which can confound genotype-phenotype correlations. To overcome these limitations, we generated a panel of MFN2-KO cells re-expressing MFN2 variants chosen to represent a variety of pathologies, which includes 6 variants linked to myopathy (Table 1) (Figure 1A). We first confirmed that re-expression levels in our U2OS MFN2 KO cells were comparable to endogenous MFN2 (Figure 1A). Next, we quantified amounts of mtDNA release via microscopy. We observed significant mtDNA release in the MFN2 -/-cells, which is almost completely rescued by re-expression of the full-length WT protein (Figure 1C-D). Surprisingly, none of the pathogenic variants completely rescued mtDNA release to the KO+WT levels (Figure 1D, Figure S1A), suggesting that all MFN2 variants investigated cause mtDNA release to varying degrees.

**Figure 1:**
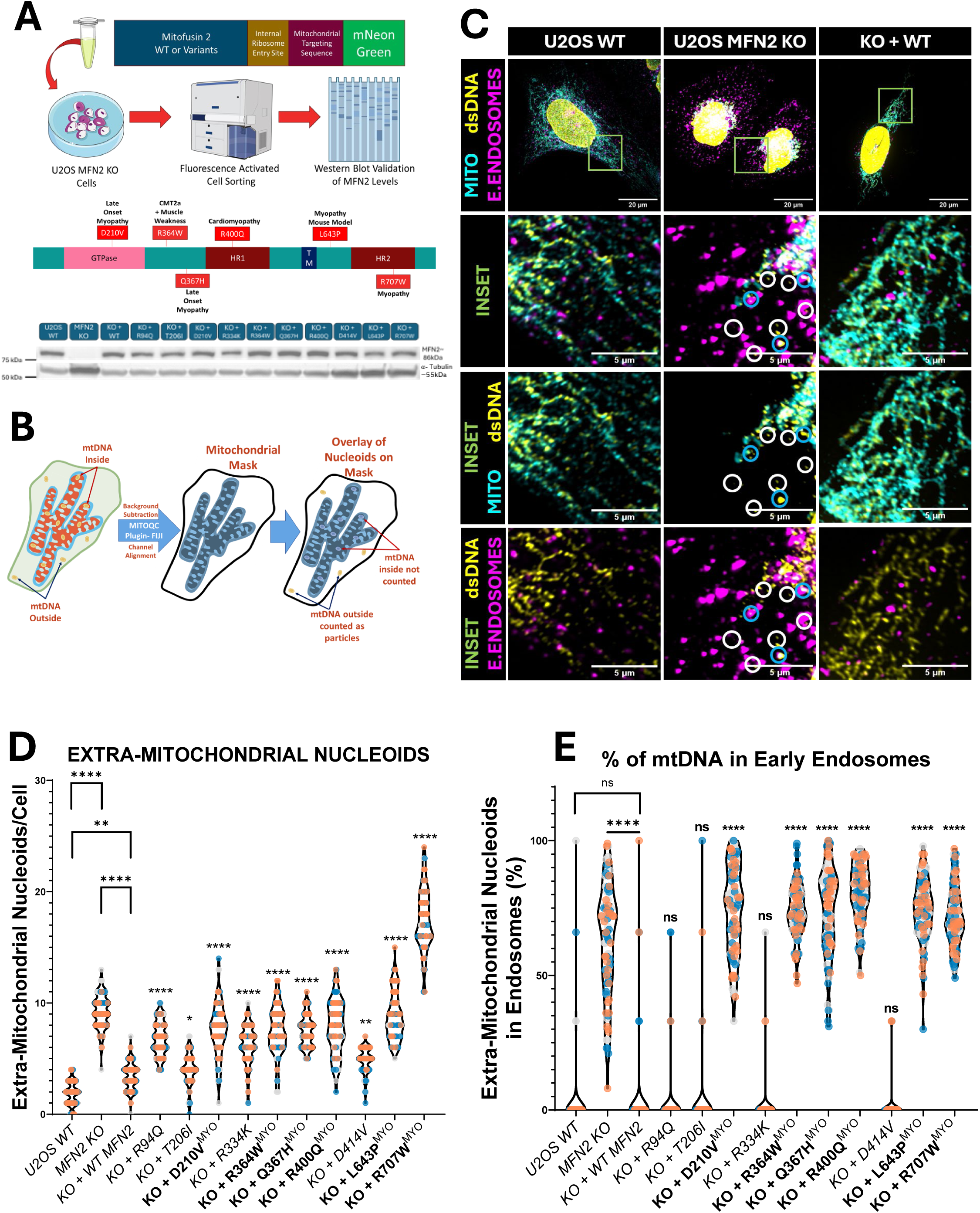
Pathogenic MFN2 variants cause mtDNA release in KO-Re-Expression cells. (A) Schematic diagram showing the process of knockout re-expression of MFN2 variants in MFN2 KO cells and a representative Western blot looking at MFN2 and Beta-Tubulin levels in KO-re-expression cells. The MFN2 variants linked to myopathy are also shown in the schematic. (B) Schematic diagram showing the process of automated quantification of extra-mitochondrial mtDNA, using the MitoQC plugin in FIJI. (C) Representative confocal images showing WT U2OS, MFN2 KO and re-expression of WT MFN2 in MFN2 KO cells, stained for mtDNA (anti-dsDNA) in yellow, mitochondria (anti-TOMM20) in cyan and early endosomes (anti-RAB5C) in magenta. The top images are cropped in the green boxes and insets are shown accordingly. The blue circles indicate extra-mitochondrial mtDNA that colocalizes with Rab5 early endosomes, while the white circles show extra-mitochondrial mtDNA that is not in Rab5 early endosomes. (D) Quantification of the number of extra-mitochondrial nucleoids per U2OS WT/MFN2 KO and re-expression variants. (E) Quantification of extra-mitochondrial mtDNA in Rab5 early endosomes as a percentage of total mtDNA outside the mitochondrial network. For D and E, the violin plots show the median and interquartile ranges; n=3, each replicate has 30 cells, the points show technical replicates and colours indicate the biological replicates. Statistics shown are one-way ANOVA; all statistics are compared to KO + WT re-expression, unless otherwise shown. Statistical symbols indicate- ns=P > 0.05, *=P ≤ 0.05, **= P ≤ 0.01, ****= P ≤ 0.0001.

**Table 1:** MFN2 variants of interest studied in this paper. The table lists all variants of interest that were studied in this paper. The table also specifies which variants are linked to myopathy, peripheral neuropathy or both and the zygosity of the patients. All variants (except the novel insertion variant) were re-expressed in U2OS MFN2 KO cells using the pipeline shown in Figure 1. The patient fibroblasts used in this study are R334K, Q367H, R397_Q398insKR, D414V and R707W.

| Variant | Domain | Muscle Pathology Reported | Peripheral Neuropathy Reported | Zygosity | Additional Pathologies |
| --- | --- | --- | --- | --- | --- |
| R94Q | GTPase | NO | YES | HET | Optic Atrophy |
| T206I | GTPase | NO | YES | HET | Cerebellar Ataxia |
| D210V | GTPase | YES | YES | HET | Optic Atrophy |
| R334K | GTPase | NO | NO | HOMO | Areflexia, Arthrogryposis Encephalopathy Hypotonia, Respiratory Failure |
| R364W | INTER-DOMAIN | YES | YES | HET | Optic Atrophy, Vocal Cord Paralysis |
| Q367H | INTER-DOMAIN | YES | NO | HET | N/A |
| R397_Q398 insKR | HR1 | YES-FIRST REPORTED HERE | NO | HET | N/A |
| R400Q | HR1 | NO | YES | HET | Cardiomyopathy |
| D414V | HR1 | NO | NO | HOMO | Cerebellar Ataxia, Optic Atrophy, Sensorineural Hearing Loss, Diabetic Neuropathy |
| L643P | INTER-DOMAIN | YES | NO | HOMO (MOUSE) | Bone Abnormalities |
| R707W | HR2 | YES | YES | HOMO | Lipodystrophy |
| R707W | HR2 | YES | YES | HET | N/A |

### MFN2 variants linked to myopathy promote mtDNA release via endosomes

Given previous findings linking MFN2 dysfunction to endosomal mtDNA release (Irazoki et al., 2023; Zaman et al., 2025), we co-stained our lines expressing MFN2 variants for mtDNA and RAB5+ve early-endosomes (Figure 1E, Figure S1D-E). Strikingly, only MFN2 variants associated with muscle pathologies showed high levels of mtDNA in RAB5+ve early-endosomes. Notably, in the cells expressing MFN2 myopathy variants, mitochondria and endosomes were in closer proximity (Figure S1G), and there were more Rab5C early endosomes, which were also enlarged (Figure S1H-I). To better understand the dynamics between mitochondria and RAB5+ early endosomes in more detail, we performed time lapse imaging of WT and MFN2 KO cells and found that KO cells had higher portion of RAB5+ early endosomes in proximity to mitochondria (Figure S1F).

### MFN2 loss leads to endosomal and MDV pools of extramitochondrial mtDNA

While we see a clear correlation between MFN2 myopathy variants and mtDNA in early endosomes, our findings raise questions about the nature of the extra-mitochondrial mtDNA for the remaining MFN2 variants, which did not co-localize with endosomes (Figure 1D, Figure S1E). To further distinguish between the two types of extra-mitochondrial mtDNA puncta, we quantified their sizes in MFN2 KO cells, which contain significant numbers of both endosomal and non-endosomal extra-mitochondrial mtDNA (Figure 2A-B). Notably, mtDNA nucleoids colocalizing with endosomes were greater than 0.3µm^2^, while non-endosomal puncta were smaller than 0.3µm^2^ (Figure 2B). Applying this size-based segregation to our panel of MFN2 variants, we again observed a correlation of ‘larger’ extra-mitochondrial mtDNA in myopathy-linked variants (Figure 2C). In contrast, non-myopathy-linked variants mainly showed ‘smaller’ released mtDNA (Figure 2D). Thus, these two pools of extra-mitochondrial mtDNA can be distinguished based on either endosomal co-localization or size.

**Figure 2:**
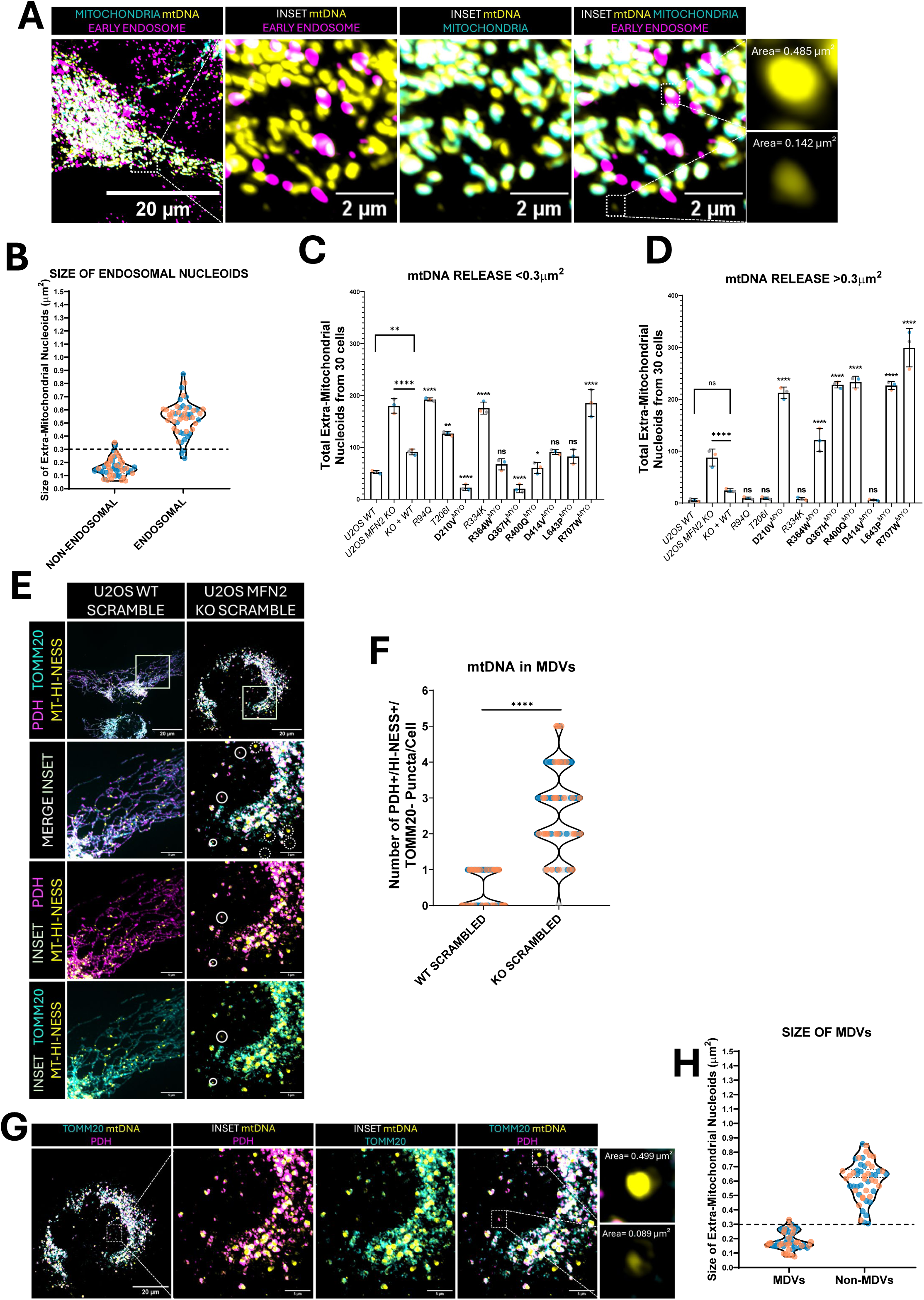
Two pools of mtDNA release in MFN2 KO cells through endosomes and MDVs. (A) Representative confocal images showing the region of U2OS MFN2 KO cells stained for mitochondria (anti-TOMM20) in cyan, mtDNA (anti-HA) and early endosomes (anti-RAB5C), where the white box in the far-left image is enlarged in the insets. The two boxes indicated in the third image, one showing mtDNA in early endosomes and one not in early endosomes are further enlarged in the fourth image. (B) Quantitative analysis of all mtDNA from 50 cells, divided into those co-localized with early endosomes and those not. The violin plots show median and IQR, with the striped line showing the cutoff for segregating the two pools of extra-mitochondrial mtDNA (endosomal vs non-endosomal), points indicate an extra-mitochondrial nucleoid and colours represent biological replicates. (C-D) Quantitative analyses of extra-mitochondrial mtDNA separated into (C) small and (D) large puncta, as per the threshold in (B). The bars in (C-D) indicate the mean +/- SD of extra-mitochondrial mtDNA, n=3, each point indicates the total extra-mitochondrial mtDNA from 30 cells, colours indicate biological replicates. (E) Representative confocal images of U2OS WT and MFN2 KO cells showing MDVs, stained for the mitochondrial matrix (anti-PDH), the outer mitochondrial membrane (anti-TOMM20) and mtDNA (anti-HA). The boxes in the top panel are enlarged in the bottom panels, the white circles indicate PDH+/mtDNA+/TOMM20- MDVs. (F) Quantitative analyses of the number of PDH+/mtDNA+/TOMM20- MDVs, n=3, each replicate has 30 cells. The violin plots show the median number of MDVs and IQR. (G) Representative confocal images showing region of U2OS MFN2 KO cells stained for MDVs, stained for the mitochondrial matrix (anti-PDH), the outer mitochondrial membrane (anti-TOMM20) and mtDNA (anti-HA). The regions with white boxes in the third image are enlarged to show the size differences in the fourth image. (H) Quantitative analysis of all extra-mitochondrial mtDNA from 50 cells, divided into those co-localized with PDH+/TOMM20- MDVs and those not. The violin plots show median and IQR, with the striped line showing the cutoff for segregating the two pools of extra-mitochondrial mtDNA (MDVs and non-MDVs), points indicate an extra-mitochondrial nucleoid and colours represent biological replicates. Statistics shown are one-way ANOVA, all statistics are compared to KO + WT re-expression, unless otherwise shown. Statistical symbols indicate- ns=P > 0.05, *=P ≤ 0.05, **= P ≤ 0.01, ****= P ≤ 0.0001.

To determine if the smaller mtDNA structures might be in PDH+/TOMM-MDVs, which have been linked to mtDNA release (Nguyen et al., 2025; Zecchini et al., 2023), next, we co-stained mtDNA in MFN2 KO cells with PDH (a matrix marker) and TOMM20 (an outer membrane marker) . We observed both a significant increase in the number of PDH+/TOMM-MDVs, as well as the presence of mtDNA within a subset of these MDVs (Figure 2E-F, Figure S2E-I). To validate that these structures were MDVs, we knocked down expression of sorting nexin 9 (*SNX9*) (Zecchini et al., 2023), which depleted the numbers of MDV puncta (Figure S2E-H). At the same time, the abundance of extramitochondrial mtDNA puncta below the 0.3µm^2^ threshold was also reduced (Figure 2G-H), further validating the size-based segregation approach for these two classes of extramitochondrial mtDNA. Ultimately, we find that MFN2 KO cells harbor two pools of extra-mitochondrial mtDNA, with larger mtDNA puncta greater than 0.3µm^2^ present in endosomes, and smaller mtDNA puncta less than 0.3µm^2^ in MDVs.

### Endosome and MDV release pathways are distinct

Next, we investigated whether the endosomal and MDV mtDNA pools in MFN2 KO cells represent independent release mechanisms, or if they are subsequent steps in a single mtDNA release pathway. To assess this idea, we either blocked endosomal mtDNA release by expressing a dominant-negative RAB5 (RAB5-DN) (Figure S2L), which prevents early endosome formation (Newman et al., 2024) (Figure S2H-J), or blocked MDVs via SNX9 depletion (Figure S2E-H). Remarkably, SNX9 knockdown decreased the abundance of mtDNA in MDVs, but slightly increased the abundance mtDNA in early endosomes (Figure 3A-B, Figure S3A). In contrast, RAB5-DN expression decreased the presence of mtDNA in early endosomes but increased the amount of mtDNA puncta in MDVs (Figure 3C-D, Figure S3B). Consistent with these findings, size segmentation analysis of ‘large’ versus ‘small’ mtDNA puncta also shows that SNX9 knockdown reduces the smaller ‘MDV’ puncta, while RAB5-DN expression reduces the larger ‘endosomal’ puncta (Figure S2C-D).

**Figure 3:**
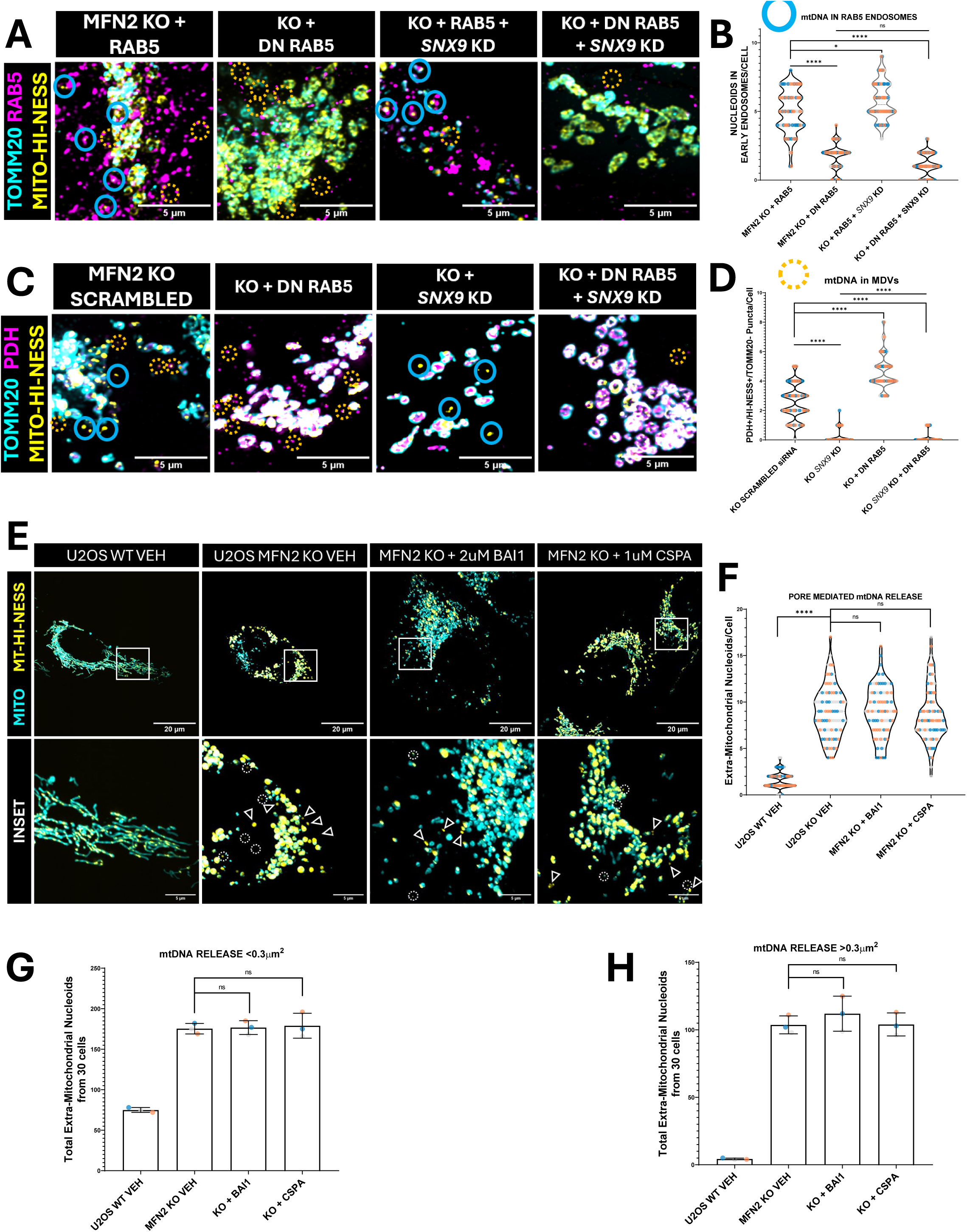
MFN2 KO cells show two distinct mechanisms of mtDNA release. (A) Representative confocal images of U2OS MFN2 KO cells with the treatments defined, stained for mitochondria (anti-TOMM20), mtDNA (anti-HA) and early endosomes (anti-RAB5C). The blue circles indicate extra-mitochondrial mtDNA that co-localizes with Rab5 early endosomes, while the yellow dotted circles indicate extra-mitochondrial mtDNA that do not. (B) Quantification of the number of endosomal mtDNA per cell, n=3, each replicate has 30 cells, each point represents a cell and colours indicate biological replicates. The violins show the median number of mtDNA and the IQR. (C) Representative confocal images of U2OS MFN2 KO cells with the treatments defined, stained for the mitochondrial matrix (anti-PDH), the outer mitochondrial membrane (anti-TOMM20) and mtDNA (anti-HA). The blue circles indicate extra-mitochondrial mtDNA that co-localizes with PDH+/TOMM20- MDVs, while the yellow dotted circles indicate extra-mitochondrial mtDNA that do not. (D) Quantification of the number of PDH+/mtDNA+/TOMM20- MDVs per cell, n=3, each replicate has 30 cells, each point represents a cell and colours indicate biological replicates. The violins show the median number of MDVs and the IQR. (E) Representative live cell confocal images of mtDNA in U2OS WT or MFN2 KO cells with the treatments defined, stained for mitochondria (MitotrackerDeepRed) and mtDNA (mt-HI-NESS). (F-H) Quantification of the (F) number of extra-mitochondrial mtDNA and the number of (G) small and (H) large mtDNA outside the mitochondrial network. The violin plots in (F) represent the median number of extra-mitochondrial mtDNA and IQR, n=3, each replicate has 30 cells, with each point showing a cell and colours indicating biological replicates. The bar graphs in (G) and (H) show the mean +/- SD of extra-mitochondrial mtDNA, n=3, each point indicates the total extra-mitochondrial mtDNA from 30 cells, and colours indicate biological replicates. Statistics shown are one-way ANOVA, with comparisons outlined. Statistical symbols indicate- ns=P > 0.05, *=P ≤ 0.05, ****= P ≤ 0.0001.

We further validated the total numbers of extra-mitochondrial mtDNA and found that blocking either endosomes or MDVs did not fully rescue the number of extra-mitochondrial mtDNA puncta. However, blocking both pathways restored mtDNA release levels to that observed in WT cells (Figure 3A-D, Figure S2A-B, Figure S3C). Consistent with the absence of any other mtDNA release mechanisms, the addition of small-molecule inhibitors of known pore-mediated mtDNA release pathways did not reduce mtDNA release in MFN2-KO cells (Fig 3 E-H). Overall, these findings demonstrate that MFN2 dysfunction causes mtDNA release through distinct endosomal and MDV pathways.

### MFN2 loss increases the frequency of mtDNA release events in live cells

Although fixed-cell imaging established that MFN2 dysfunction promotes the accumulation of extra-mitochondrial mtDNA, this endpoint approach does not capture the kinetics of release or resolve how individual mtDNA structures emerge and persist over time. We therefore developed and applied a live-cell mtDNA release tracking pipeline to directly quantify release as a dynamic process in WT and MFN2 KO cells. This approach allowed us to measure the appearance of newly released mtDNA structures, and their cumulative accumulation over time, thereby distinguishing active release behaviour from endpoint accumulation. Time-lapse imaging revealed discrete mtDNA release events emerging outside the mitochondrial network in MFN2 KO cells, with significantly greater cumulative release over time compared with WT controls (Figure 4).

**Figure 4:**
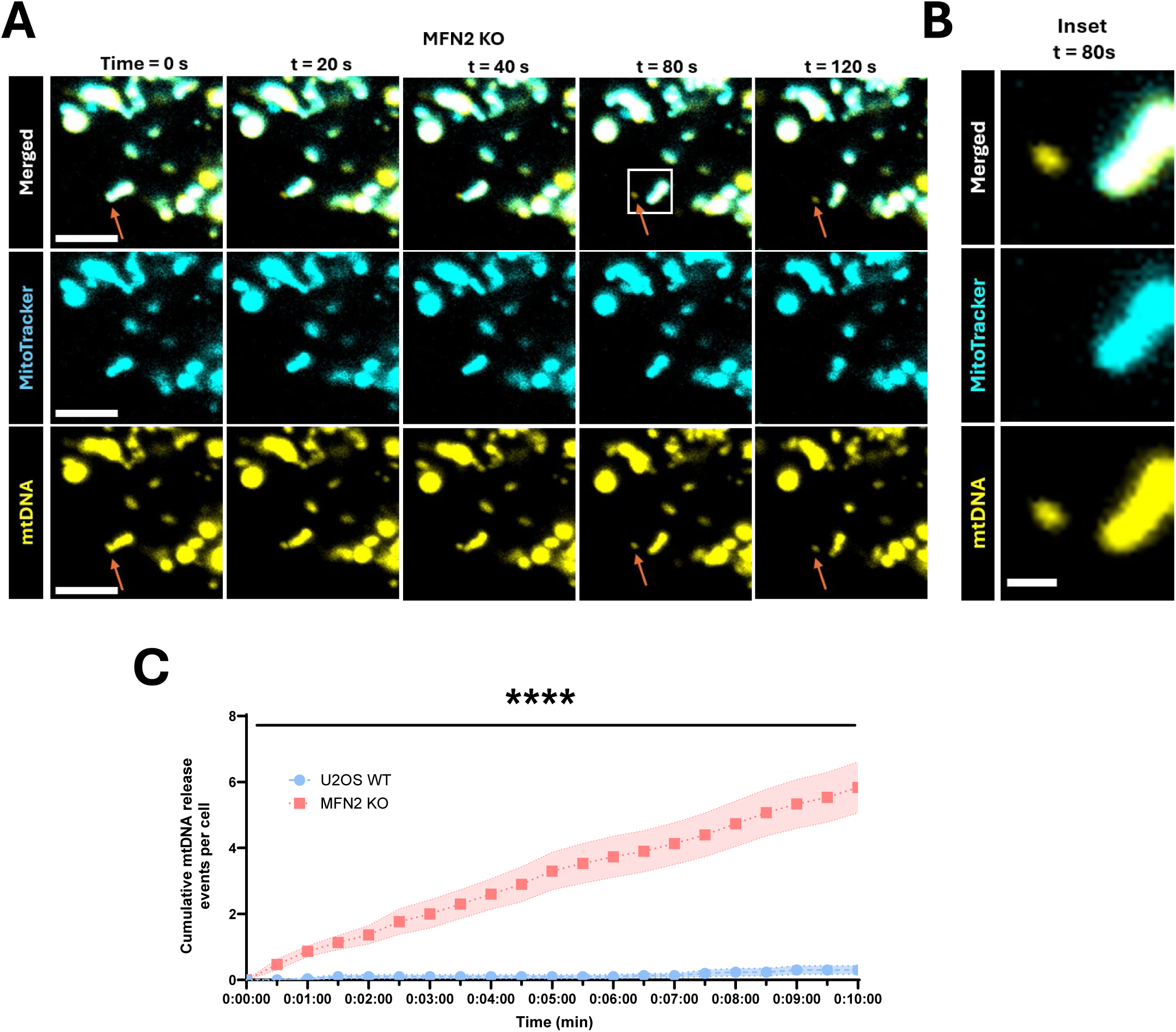
Live-cell imaging reveals temporal mtDNA release and dynamic mitochondria– Rab5+ endosome interactions in MFN2-deficient cells. (A) Representative time-lapse confocal images showing a region of U2OS MFN2 KO cells exhibiting mtDNA release over time. Mitochondria shown in Magenta were labelled with MitoView Green and mtDNA (Mt-HI-NESS). (B) Enlarged region of interest showing spatial separation between mitochondria and mtDNA at timepoint 80. (C) Quantification of cumulative mtDNA release events per cell over a 10 min imaging period in WT and MFN2 KO cells, imaged at 30 s intervals. Data are presented as mean ± SEM from three independent experiments (n = 3; 10 cells per experiment). Statistics indicate Two-way repeated-measures ANOVA with Geisser–Greenhouse correction revealed a significant genotype × time interaction (**** P < 0.0001), indicating that mtDNA release accumulated differently over time in WT and MFN2 KO cells.

### ISR activation reduces levels of extra-mitochondrial mtDNA

Given previous work showing that ISR activation can rescue multiple types of MFN2 dysfunction (*i.e.*, impaired fusion, altered MERCs, and reduced respiration) (Baron et al., 2025; Bora et al., 2025; Zaman et al., 2026), we wanted to test if this approach could also rescue mtDNA release in our MFN2 KO cells. To this end, we treated cells with Halofuginone, a prolyl-tRNA synthetase inhibitor that activates the ISR through the GCN2 arm, or with Parogrelil, which activates the ISR through the HRI axis (Hori et al., 2009). With both drugs, the number of extra-mitochondrial mtDNA was reduced after 6 hours of treatment. This rescue was blocked by cotreatment with the ISR inhibitor ISRIB, indicating the rescue is dependent on ISR activation (Figure 5A-D, Figure S4A-B). Notably, both the ‘smaller’ and ‘larger’ extra-mitochondrial mtDNA puncta were reduced, suggesting ISR activation impacts both endosome and MDV mechanisms of mtDNA release (Figure 5C-D).

**Figure 5:**
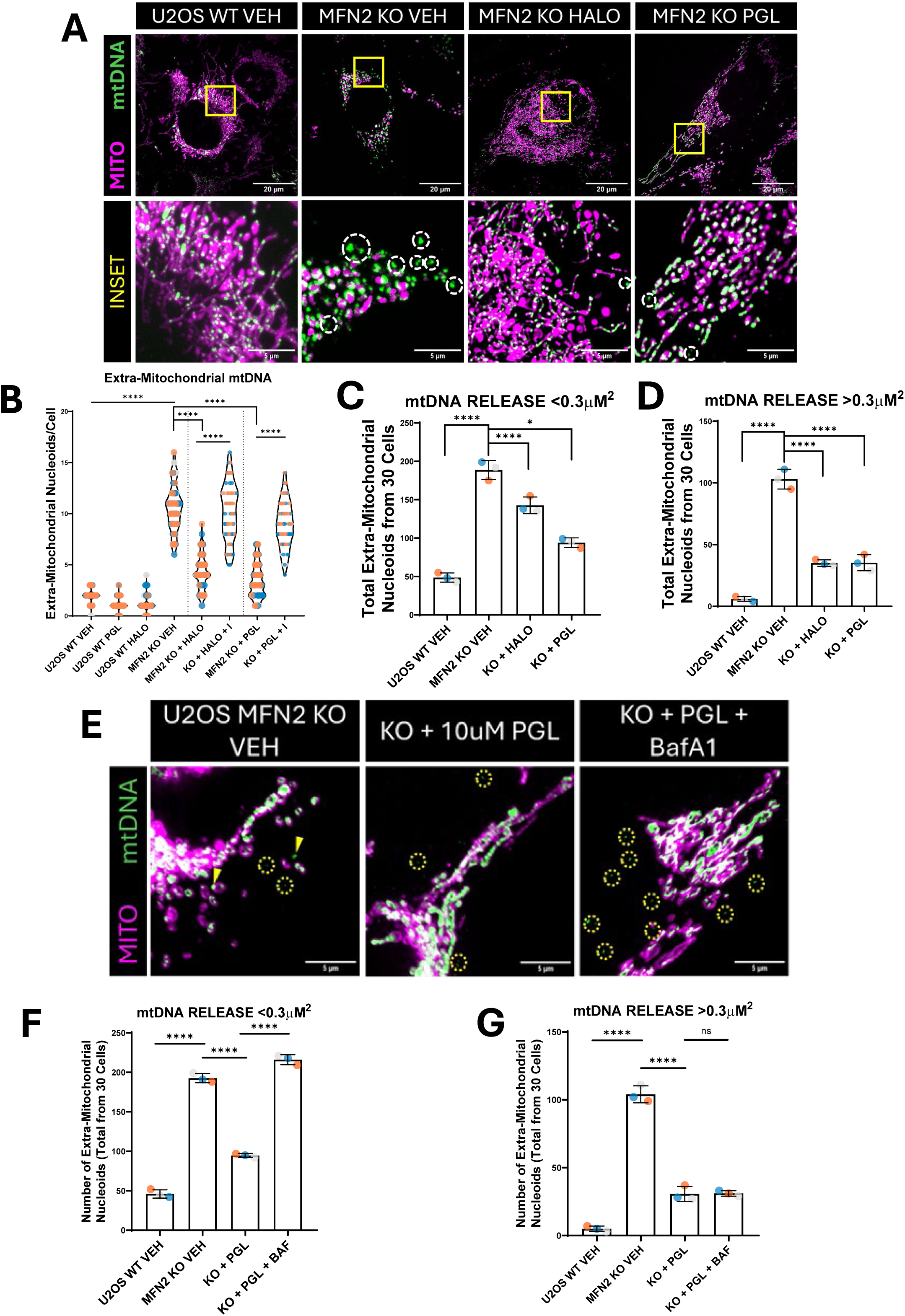
ISR activation ameliorates extra-mitochondrial mtDNA. (A) Representative live-cell confocal images showing extra-mitochondrial mtDNA in U2OS WT/MFN2KO cells and outlined treatments, stained for mitochondria (MitotrackerDeepRed) and mtDNA (mt-HI-NESS). The yellow boxes are enlarged in the bottom panel and the white circles show mtDNA outside the mitochondrial network. (B-D) Quantitative analyses of mtDNA release looking at (B) total extra-mitochondrial mtDNA and (C) small vs (D) large mtDNA according to our size segmentation of MDVs vs Endosomal mtDNA, respectively. The violin plots in (B) represent the median number of extra-mitochondrial mtDNA and IQR, n=3, with each replicate has 30 cells, with each point showing a cell and colours indicating biological replicates. The bar graphs in (C) and (D) show the mean +/- SD of extra-mitochondrial mtDNA, n=3, each point indicates the total extra-mitochondrial mtDNA from 30 cells, colours indicate biological replicates. (E) Representative live-cell confocal images looking at U2OS MFN2 KO cells and outlined treatments, stained for mitochondria (MitotrackerDeepRed) and mtDNA (mt-HI-NESS). The yellow circles indicate extra-mitochondrial mtDNA. (F-G) Quantitative analysis of (F) small vs (G) large extra-mitochondrial mtDNA, according to our sizing pipeline. The bars in (F) and (G) show the mean +/- SD of extra-mitochondrial mtDNA, n=3, each point indicates the total extra-mitochondrial mtDNA from 30 cells, colours indicate biological replicates. All statistics shown are unpaired t-tests, statistical symbols indicate- ns=P > 0.05, *=P ≤ 0.05, ****= P ≤ 0.0001.

Next, we investigated how ISR activation reduced the levels of extra-mitochondrial DNA. We reasoned that ISR activation was either preventing the release of mtDNA or increasing its turnover. As both endosome and MDV pathways are thought to deliver mtDNA to lysosomes, we tested whether ISR activation increased the degradation of extra-mitochondrial mtDNA via lysosomes. To this end, cells were treated with both ISR activators and the lysosomal inhibitor bafilomycin (Figure 5 E-G, Figure S5E-H). Notably, bafilomycin treatment abolishes the ISR-mediated rescue of release of small mtDNA puncta via MDVs. However, ISR treatment along with Bafilomycin did not impact the rescue of larger mtDNA puncta in endosomes. These findings suggest that ISR activation reduces mtDNA in MDVs by increasing their lysosomal turnover, but reduces mtDNA in endosomes by preventing their release. Overall, these findings promote ISR activation as a novel approach to mitigate mtDNA release in the context of MFN2 dysfunction.

### MFN2 patient cells linked to myopathy show mtDNA release and inflammation

While the KO-re-expression approach allows direct comparison of multiple MFN2 variants in an isogenic background and avoids the challenge of obtaining samples from rare patients, it does have limitations. For example, many pathogenic MFN2 variants are heterozygous and, as such, are not fully reflected by a KO-re-expression approach. In addition, U2OS cells are immortalized and do not have fully active inflammatory pathways that allow us to examine the downstream consequences of mtDNA release. To address these limitations, we examined mtDNA release and downstream inflammation in a subset of fibroblasts from patients with MFN2 variants to which we had access, including: R334K, Q367H, D414V, and R707W.

In addition to these fibroblasts harboring previously described MFN2 variants (Sawyer et al., 2015; Sharma et al., 2022; Zaman et al., 2025, 2026), we obtained fibroblasts from a patient with a novel heterozygous variant in MFN2, R397_insKR (c.1191_1192insAAGCGG p.Arg397_Gln398insLysArg) who had a late-onset clinical presentation featuring combined myopathy and neuropathy. This male individual first noted symptoms of muscle weakness at age 60-65 which gradually progressed, and predominantly affected distal leg musculature. Clinical examination revealed symmetric weakness most notable in ankle plantarflexion and hallux extension (MRC grade 4) with minor weakness of hip flexion/abduction, knee flexion and ankle plantarflexion (MRC grade 4+). There was no *pes cavus*, hammer toes, or intrinsic foot muscle atrophy; no sensory abnormalities were present. CK enzyme levels were elevated (597 to 837 U/L, normal range <350 U/L), including at measurements up to 8 years prior to symptom onset. Neurophysiology and EMG showed a mixture of neurogenic motor and myopathic findings. Similarly, a *vastus lateralis* muscle biopsy at age 70-75 showed nonspecific changes of a chronic, non-immune mediated, mildly active myopathy, and neurogenic changes. Genetic testing included a 330 gene panel encompassing genetic causes of myopathy and neuropathy. This identified a novel heterozygous variant of uncertain significance in *MFN2*, c.1191_1192insAAGCGG, predicted to cause p.(Arg397_Gln398insLysArg). No pathogenic variants were identified, nor were there any other variants of unknown significance considered to be potentially clinically relevant. Parents were not available for testing and were not known to have any neurologic disease. No other family members were clinically affected by any neurologic condition at the time of last assessment. To our knowledge, this is the first insertion variant found linked to MFN2 pathology. Relative to control fibroblasts, R397_insKR patient cells showed fragmentation of the mitochondrial network morphology (Figure S6C) and a 20% reduction in mtDNA copy number (Figure S6D), features consistent with MFN2 dysfunction. Given that this patient exhibited a myopathy phenotype, we also included it among our panel of MFN2 variant fibroblasts, predicting that it would exhibit endosomal mtDNA release.

When interrogated for mtDNA release, all MFN2 variant fibroblasts displayed elevated mtDNA release compared to control fibroblasts (Figure 6A-B), resembling the U2OS KO-re-expression findings above. Furthermore, fibroblasts from patients with myopathy (Q367H, R707W, R397_insKR) exhibited extra-mitochondrial mtDNA in early endosomes, reduced distances between mitochondria and early endosomes, and a greater number of endosomes, which are also enlarged (Figure S6H-J). Meanwhile, in fibroblasts from patients without myopathy (R334K & D414V), the extra-mitochondrial mtDNA was not in endosomes (Figure 6C-D, Figure S6F-G).

**Figure 6:**
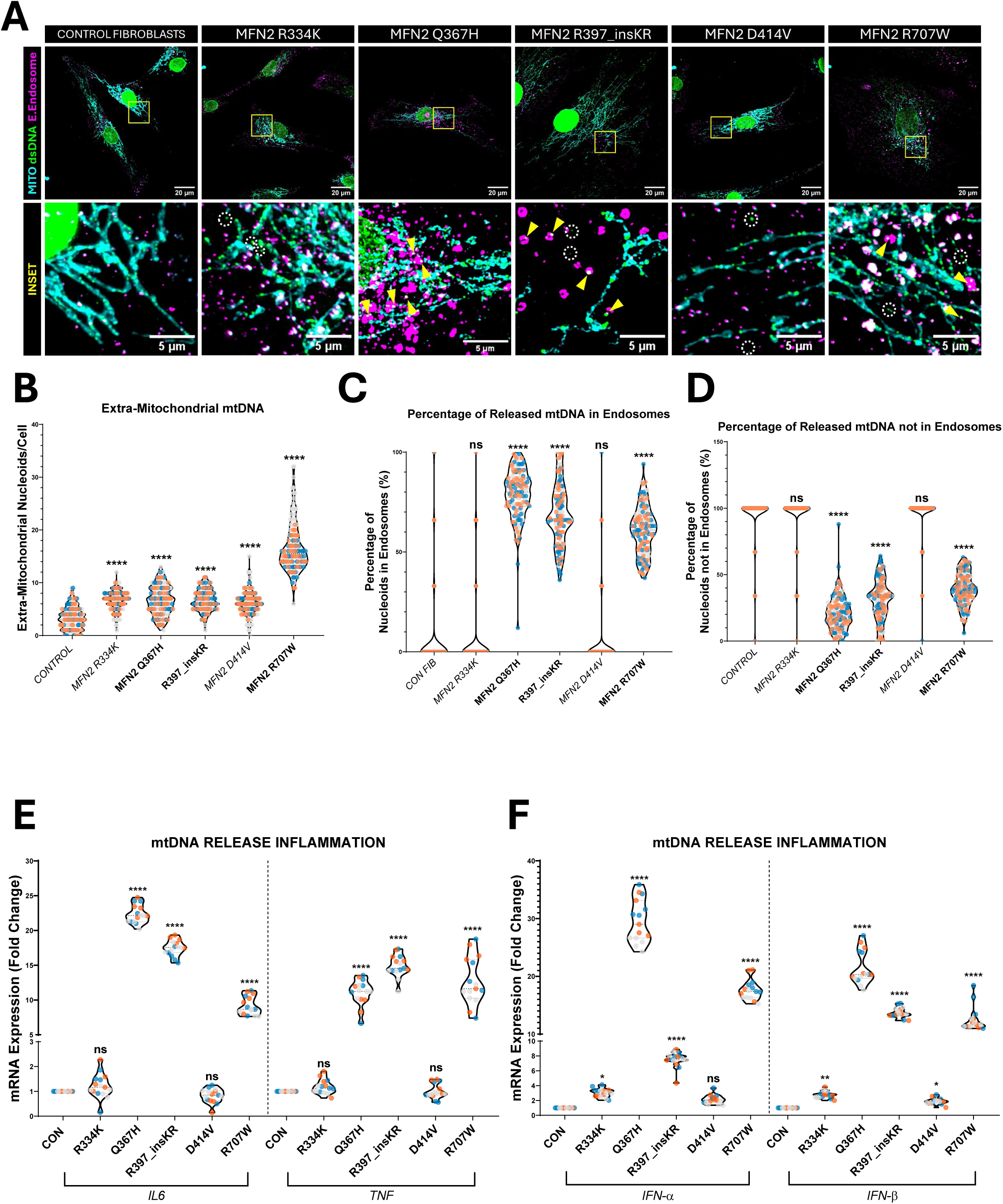
Pathogenic MFN2 variants show mtDNA release and myopathy-linked variants show inflammation. (A) Representative confocal images showing control fibroblasts from a healthy donor and patient fibroblasts from patients with the MFN2 variants outlined, stained for mtDNA (anti- dsDNA) in yellow, mitochondria (anti-TOMM20) in cyan and early endosomes (anti-RAB5C) in magenta. The top images are cropped in the yellow boxes and insets are shown accordingly. The yellow triangles indicate extra-mitochondrial mtDNA that colocalizes with Rab5 early endosomes, while the white circles show extra-mitochondrial mtDNA that is not in Rab5 early endosomes. (B-D) Quantification of the (B) total number of extra-mitochondrial nucleoids, (C) percentage of mtDNA in endosomes and (D) percentage not in endosomes. The violin plots represent the median and IQR, n=3, each point indicates the total extra-mitochondrial mtDNA from 150 cells, colours indicate biological replicates. (E-F) Quantitative analysis of relative mRNA expression of cytokines in the (E) TLR9-NF-kB pathway and (F) cGAS-STING pathway. The violin plots in (E-F) represent the median and IQR values of mRNA expression, n=3, each with 5 technical replicates. All statistics shown are one-way ANOVA comparing to the control fibroblasts, unless otherwise shown. The statistical symbols indicate- ns=P > 0.05, *=P ≤ 0.05, **= P ≤ 0.01, ****= P ≤ 0.0001.

Finally, we measured the expression of inflammatory markers in the patient fibroblast lines. As general markers of TLR9 activation, we examined *TNF-*α and *IL-6* (Fang et al., 2016; Irazoki et al., 2023; Zaman et al., 2025). In parallel, we looked at levels of *Interferon-* α*/*β as reporters of cGAS-STING signaling (Al Khatib et al., 2023; Irazoki et al., 2023; Zaman et al., 2025). Consistent with the mtDNA release patterns observed by microscopy, the myopathy-linked MFN2 variants show strong (∼8-30 fold) activation of both TLR9 and cGAS-STING inflammatory markers. In contrast, the other MFN2 variants show only a modest (2-3 fold), if any, increase of cGAS-STING inflammatory markers, and no indication of TLR9 activation. Altogether, these findings point to myopathy-linked MFN2 variants promoting mtDNA release predominantly through the endosomal pathway, leading to downstream TLR9 and cGAS-STING activation and inflammation.

## Discussion

While considerable attention has been focused on mechanisms underlying CMT2A neuropathy, other MFN2-associated pathologies (*e.g.*, myopathy, cerebellar ataxia, lipomatosis) have largely been ignored with respect to their underlying mechanisms. Here, we identify endosomal mtDNA release and TLR9-mediated inflammation as conserved hallmarks of MFN2-myopathies. More broadly, our findings reveal that MFN2 dysfunction promotes mtDNA release through two mechanistically distinct vesicular release pathways, with different inflammatory consequences, thus positioning mtDNA release as a previously unrecognized determinant of MFN2 disease heterogeneity.

A central advance made in this study is the identification of endosomal mtDNA release as a conserved feature of MFN2 variants linked to myopathy. Across both isogenic cell models and patient-derived fibroblasts, only myopathy-associated variants (including the novel R397_inskR variant) display mtDNA in RAB5-positive compartments. This early-endosome mtDNA release phenotype was accompanied by enhanced inflammatory signaling in patient fibroblasts. Together with previous reports showing TLR9 inflammation is sufficient to drive muscle inflammation and pathology (Brunn et al., 2012; Rodríguez-Nuevo et al., 2018), these findings support a model in which aberrant endosomal mtDNA release contributes to the pathogenesis of MFN2-myopathies.

Myopathy as a standalone finding in MFN2 dysfunction has important clinical implications. While myopathy and muscle weakness has long been recognized as a phenotype in a subset of CMT2a patients (Banchs et al., 2009; Rouzier et al., 2012; Stuppia et al., 2015; Dorn, 2020), it can also occur in the absence of peripheral neuropathy (Zaman et al., 2025), a finding recapitulated in a mouse model of MFN2 dysfunction (Hines et al., 2023). The clear mechanistic correlation between myopathy and TLR9 inflammation further supports the notion that myopathy can occur independent of neuropathy. Therefore, *MFN2* should be thought of as a candidate pathogenic gene with the potential to cause myopathy in the clinic, not just peripheral neuropathy. The novel case we describe here (with the R397inKR variant) supports this notion, but it remains possible that additional genetic variants not identified by the 330 gene panel could be contributing to the patient phenotype. Because the variant is present at very low frequency in population databases, it may represent a reduced penetrance variant, with a phenotype easily overlooked in late-onset clinical presentations. Identification of additional unrelated individuals with this variant and a myopathy/neuropathy phenotype will help provide support for the pathogenicity of this variant.

Our findings may also have broader implications for inflammation in MFN2 disease beyond myopathy, as non-myopathy MFN2 variants displayed non-endosomal mtDNA release and modest activation of cGAS-STING inflammation. Although the mechanisms underlying MFN2-induced peripheral neuropathy are incompletely understood, growing evidence implicates cGAS-STING-dependent neuroinflammation in both central and peripheral nervous system pathologies (Gan et al., 2026; Jiang et al., 2026; Liu et al., 2024; Zhang et al., 2025). The modest cGAS-STING activation observed in non-myopathic MFN2 variants therefore raises the possibility that chronic mtDNA release-driven inflammatory signalling contributes to the neuropathy pathogenesis. It will therefore be important to determine whether chronic mtDNA release and inflammation contribute to MFN2-mediated peripheral neuropathy.

An unexpected mechanistic advance from this research is that MFN2 dysfunction simultaneously engages two mechanistically distinct vesicular mtDNA release pathways. Previous studies have described mtDNA release through either RAB5-positive compartments (Newman et al., 2024) or MDVs (Nguyen et al., 2025; Zecchini et al., 2023). As these pathways have been studied under different biological contexts, it is unclear whether they represent independent mechanisms or sequential stages of a common route. For example, mtDNA could be routed towards early endosomes via MDVs. However, previous reports show that mtDNA-containing MDVs are destined for late endosomes/lysosomes (Nguyen et al., 2025). Meanwhile, several aspects of our findings support the notion that MDVs and endosomes are independent pathways of mtDNA release. First, genetic disruption of RAB5-dependent endosomal release selectively disrupts endosomal mtDNA, without diminishing mtDNA in MDVs. Second, *SNX9* depletion selectively impaired MDV formation without reducing endosomal mtDNA. Third, blocking either pathway seems to reciprocally upregulate the other pathway, suggesting compensatory mtDNA release mechanisms, which would not be seen if they were part the same route. Fourth, pharmacological ISR activation affects endosomal and MDV mtDNA release differently. Specifically, our data show that ISR activation blocks endosomal packaging of mtDNA, while simultaneously increasing the turnover of mtDNA in MDVs. Fifth, the fact that only MFN2 variants linked to myopathy show endosomal release, while other variants exhibit MDV release also supports the notion that these pathways are distinct. Finally, the increased interactions between mitochondria and early endosomes, which is also reported in the context of impaired MFN2 function by other groups (Das et al., 2016; Irazoki et al., 2023), argues against the need for MDVs to deliver mtDNA to endosomes. Collectively, these observations suggest that dysfunctional mitochondria can engage in multiple parallel mtDNA release pathways rather than a single linear release cascade.

Our findings also extend previous observations of MDVs containing mtDNA. Although past studies have not linked MFN2 to MDV formation, similar to reports in fumarate-hydratase-deficient cells (Zecchini et al., 2023), we observed mtDNA in PDH+/TOMM20- MDVs, which require SNX9 for their formation. However, MFN2 dysfunction represents a mechanistically distinct context in which this mtDNA release pathway is engaged. These results broaden the range of mitochondrial stresses capable of activating mtDNA-containing MDV formation

An intriguing question that remains is why cells maintain independent mtDNA release pathways. One possible explanation is that these pathways are activated through different trigger/signals. In this sense, endosomal mtDNA release has been linked to mtDNA replication stress (Newman et al., 2024), whereas mtDNA present in MDVs has been explained in the context of altered mitochondrial metabolic dysfunction (Zecchini et al., 2023) or inflammatory cell death (Nguyen et al., 2025). The different sizes of the mtDNA in MDVs and early endosomes may also reflect distinct triggers. In this regard, the larger size of mtDNA in endosomes may indicate release of intact mtDNA nucleoids, while MDVs may carry mtDNA fragments or sub-nucleoid structures. Future work will need to determine how different stresses, cargo selection, and release routes all shape the fate of released mtDNA. In this regard, studying the specific pathogenic variants in more detail may help to further tease apart how MFN2 differentially impacts these release pathways.

The existence of parallel mtDNA release pathways has distinct implications for downstream inflammatory signaling. Both endosomal and MDV-associated mtDNA release appear to be routed ultimately towards lysosomal degradation (Newman et al., 2024; Nguyen et al., 2025; Zecchini et al., 2023), which can release mtDNA into the cytosol to activate cGAS-STING inflammation (Nguyen et al., 2025). Meanwhile, endosomal release uniquely positions mtDNA where it can activate TLR9 signaling. Therefore, endosomal mtDNA can activate both TLR9 and cGAS-STING inflammation. Consistent with this model, myopathy-linked MFN2 variants activate both TLR9- and cGAS-STING-associated inflammatory programs. In contrast, MDV-associated mtDNA appears to be specifically linked to cGAS-STING inflammation, as seen by others (Nguyen et al., 2025; Zecchini et al., 2023). Therefore, we propose that the intracellular route taken by released mtDNA functions as an important layer of innate immune activation.

The precise mechanism by which mtDNA enters RAB5 positive early endosomes, remains an unresolved issue in the field. However, our findings imply that MFN2 is a candidate regulator of this process. In this regard, MFN2 links mitochondria to other organelles such as the ER (de Brito & Scorrano, 2008) and lipid droplets (Boutant et al., 2017) and has recently been linked to contacts between mitochondria and early endosomes (Gordaliza-Alaguero et al., 2025; Irazoki et al., 2023). Consistent with having a regulatory role in maintaining homeostasis of mito-endosome contact sites, MFN2 dysfunction reduced the distance between mitochondria and early endosomes and selectively promoted endosomal mtDNA release into endosomes in the myopathy-linked variants. These findings raise the possibility that altered mito-endosome interactions may facilitate direct release of mtDNA into the endosomal system. This model is consistent with our findings that PDH+/TOMM20- MDVs are not necessary for endosomal mtDNA accumulation, arguing against a simple MDV-to-endosome transfer model. More broadly, emerging work has implicated RAB5-positive endosomes in mitochondrial quality control pathways (Ravindran et al., 2022; Todkar et al., 2019) and mitophagy (Hammerling et al., 2017), suggesting endosomes may have a more direct role in mitochondrial homeostasis. Although we do not fully understand the mechanisms of mtDNA release into early endosomes, MFN2 likely plays a crucial role in gatekeeping the process.

Armed with the novel mechanistic insight that MFN2-linked myopathy is linked to TLR9 sensing of released mtDNA, it may now be possible to target this pathway to treat myopathy specifically. A general approach to treating mtDNA-mediated inflammation is to dampen the inflammation. An exciting example is the prevention of myopathy in the muscle-specific MFN1 deletion mouse by treatment with the NF-kB inhibitor sodium salicylate (Irazoki et al., 2023). Consistent with this finding, other studies also show that blocking TLR9-mediated inflammation can be beneficial in treating myopathy (Irazoki et al., 2023; Oka et al., 2012; Tripathi et al., 2023).

As an alternative to dampening inflammation, it may also be possible to prevent mtDNA release before it induces inflammation. In this regard, we found that ISR activation reduced both endosomal and MDV-associated mtDNA pools, extending our previous findings that ISR can rescue MFN2 dysfunction (Baron et al., 2025; Bora et al., 2025). Intriguingly, ISR activation reduced endosomal release events, and increased lysosomal turnover of mtDNA from MDVs. Therefore, blocking inflammation through anti-inflammatories or reducing mtDNA release through ISR activation warrants further investigation as potential remedies for MFN2-associated myopathy. Beyond MFN2 myopathy, ISR activation may also be a novel therapeutic strategy for other inflammatory diseases linked to mtDNA release.

Collectively, our findings position MFN2 as a crucial regulator of mtDNA release and inflammatory signaling. Disease-associated MFN2 variants disrupt this gatekeeping function in distinct ways, generating alternative mtDNA release routes that engage different inflammatory pathways and correlate with distinct clinical phenotypes. The strong association between endosomal mtDNA release and myopathy-linked MFN2 variants identifies the mitochondria-endosome axis as a candidate pathomechanism underlying MFN2-associated myopathy.

## Data Availability

All data produced in the present work are contained in the manuscript

## Acknowledgments

We want to thank Dr. Johnathan Canton (University of Calgary) for kindly providing the RAB5 plasmid. We want to thank Dr. Edward Fon (McGill University) for the U2OS cells. We thank the Care4Rare Canada Consortium for access to cell line carrying the MFN2 R707W variant.

## Author contributions

Designed, performed or analysed experiments: MZ, JD, AM, CC, JT, RLW, PC, GP, TS. Supervised trainees: CNA, PC, GP, TS. Clinical evaluation of patients and patient cells: GP. Drafted manuscript and figures: MZ, JD, GP, TS. All authors discussed, commented on and consented authorship on the manuscript.

## Supplementary Figure Legends

**Supplementary Figure S1: Pathogenic MFN2 variants cause mtDNA release in KO-Re-Expression cells** (A) Representative confocal images showing WT U2OS, MFN2 KO and re-expression of WT MFN2 in MFN2 KO cells, stained for mtDNA (anti-dsDNA) in yellow, mitochondria (anti-TOMM20) in cyan and early endosomes (anti-RAB5C) in magenta. The top images are cropped in the green boxes and insets are shown accordingly. The blue circles indicate extra-mitochondrial mtDNA that colocalizes with Rab5 early endosomes, while the white circles show extra-mitochondrial mtDNA that are not in Rab5 early endosomes. (B-I) Quantitative analyses of (B) numbers and (C) sizes of mtDNA nucleoids, number of extra-mitochondrial mtDNA puncta (D) co-localized with and (E) not co-localized with Rab5 early endosomes, (F) representative confocal images of mitochondria-RAB5 early endosome proximity in WT U2OS and MFN2 KO cells labelled with MitoView Green (mitochondria) and Rab5-mCherry (early endosomes), (G) average distances between mitochondria and early endosomes and (H) numbers and (I) sizes of early endosomes per cell in U2OS WT/MFN2 KO and KO re-expression variants. For (B-H), the violin plots show the median and interquartile ranges; n=3, each replicate has 30 cells, the points show technical replicates and colours indicate the biological replicates. Statistics shown are one-way ANOVA, all statistics are compared to KO + WT re-expression, unless otherwise shown. Statistical symbols indicate- ns=P > 0.05, *=P ≤ 0.05, **= P ≤ 0.01, ***= P ≤ 0.001, ****= P ≤ 0.0001.

**Supplementary Figure S2: MFN2 KO cells show two distinct mechanisms of mtDNA release** (A) Representative confocal images showing mtDNA release in U2OS WT/MFN2 KO and outlined treatments of MFN2 KO cells, stained for mitochondria (anti-TOMM20) and mtDNA (anti-HA). The yellow boxed regions are enlarged in the bottom panels, extra-mitochondrial nucleoids above 0.3µm^2^ are shown with the yellow arrows, while those below 0.3µm^2^ are shown with the yellow circles. (B-D) Quantitative analysis of the effect of *SNX9* knockdown, dominant-negative Rab5 or both on (B) mtDNA release and the pools of mtDNA release (C) small vs (D) large, according to our size segmentation of MDVs vs Endosomal mtDNA, respectively. The violin plots in (B) represent the median number of extra-mitochondrial mtDNA and IQR, n=3, with each replicate having 30 cells, with each point showing a cell and colours indicating biological replicates. The bar graphs in (C) and (D) show the mean +/- SD of extra-mitochondrial mtDNA, n=3, each point indicates the total extra-mitochondrial mtDNA from 30 cells, colours indicate biological replicates. (E) Representative confocal images in U2OS WT/MFN2 KO and respective treatments to look for MDVs stained for the mitochondrial matrix (anti-PDH), the outer mitochondrial membrane (anti-TOMM20) and mtDNA (anti-HA). The solid circles represent PDH+/TOMM20- MDVS, while the dotted circles represent extra-mitochondrial mtDNA not in MDVs. (F) Representative western blot image showing cell lysates probed for SNX9 and Beta-Tubulin for U2OS WT/MFN2 KO cells with the treatments described. (G) Quantitative analyses of the number of PDH+/mtDNA+/TOMM20- MDVs, n=3, each replicate has 30 cells. (H) Quantitative analyses of the number of PDH+/TOMM20-/mtDNA- MDVs, n=3, each replicate has 30 cells. (I) Quantitative analyses of the number of TOMM20+/mtDNA-/PDH- MDVs, n=3, each replicate has 30 cells. The violin plots in (G-I) show the median number of MDVs and IQR. (J) Representative confocal images showing mtDNA release into Rab5 early endosomes in U2OS WT/MFN2 KO and outlined treatments, stained for mitochondria (anti-TOMM20), mtDNA (anti-HA) and early endosomes (anti-Rab5C). (K) Quantitative analysis of mtDNA in early endosomes for U2OS WT/MFN2KO and outlined treatments. The violin plots show the median number of extra-mitochondrial mtDNA and IQR, n=3, each replicate has 30 cells, with each point showing a cell and colours indicating biological replicates. (L) Representative confocal images showing mtDNA release into Rab7 late endosomes in MFN2 KO cells with the outlined treatments, stained for mitochondria (anti-TOMM20), mtDNA (anti-HA) and late-endosomes (anti-Rab7). The yellow circles represent extra-mitochondrial mtDNA, while the yellow triangles show mtDNA co-localized with Rab7 late endosomes. All statistics shown are unpaired t-tests as outlined, statistical symbols indicate- ns=P > 0.05, *=P ≤ 0.05, ***= P ≤ 0.001, ****= P ≤ 0.0001.

**Supplementary Figure S3: MFN2 KO cells show two distinct mechanisms of mtDNA release** (A) Representative confocal images showing mtDNA in MDVs in U2OS WT/MFN2 KO and outlined treatments, stained for the mitochondrial matrix (anti-PDH), the outer mitochondrial membrane (anti-TOMM20) and mtDNA (anti-HA). The green triangles represent mtDNA that colocalizes with PDH+/TOMM20- MDVs, while the white circles represent extra-mitochondrial mtDNA not in MDVs. (B) Representative confocal images showing mtDNA in Rab5 early endosomes in U2OS WT/MFN2 KO and outlined treatments, stained for mitochondria (anti-TOMM20), mtDNA (anti-HA) and early endosomes (anti-RAB5C). The green triangles represent mtDNA that colocalizes with Rab5-early endosomes, while the white circles represent extra-mitochondrial mtDNA not in early endosomes. (C) Quantitative analysis of number of extra-mitochondrial mtDNA puncta in U2OS WT/MFN2 KO and outlined treatments. The violin plots show the median number of extra-mitochondrial mtDNA and IQR, n=3, with each replicate has 30 cells, with each point showing a cell and colours indicating biological replicates. All WT groups are not statistically significant, while the MFN2 KO cells show statistical significance for multiple unpaired t-tests. The statistical symbols indicate- ns=P > 0.05, ****= P ≤ 0.0001.

**Figure S4: Schematic of pipeline for quantifying kinetics of mtDNA release.**

**Figure S5: ISR activation ameliorates extra-mitochondrial mtDNA** (A-B) Representative live-cell confocal images showing extra-mitochondrial mtDNA in (A) U2OS WT and (B) MFN2 KO cells with the treatments outlined, stained for mitochondria (MitotrackerDeepRed) and mtDNA (mt-HI-NESS). (C) Representative live-cell confocal images of U2OS MFN2 KO cells and outlined treatments, stained for mitochondria (MitotrackerDeepRed) and mtDNA (mt-HI-NESS). The yellow circles indicate extra-mitochondrial mtDNA. (D-F) Quantitative analysis of (D) total number of extra-mitochondrial mtDNA per cell and (E) small vs (F) large extra-mitochondrial mtDNA, according to our sizing pipeline. The violin plots in (D) indicate the median number of extra-mitochondrial nucleoids per cell and IQR, n=3, with each replicate has 30 cells, with each point showing a cell and colours indicating biological replicates. The bars in (E) and (F) show the mean +/- SD of extra-mitochondrial mtDNA, n=3, each point indicates the total extra-mitochondrial mtDNA from 30 cells, colours indicate biological replicates. All statistics shown are unpaired t-tests, statistical symbols indicate- ns=P > 0.05, **= P ≤ 0.01, ****= P ≤ 0.0001.

**Figure S6: Pathogenic MFN2 variants show mtDNA release and myopathy-linked variants show inflammation** (A) Representative confocal images showing control fibroblasts from a healthy donor and patient fibroblasts from patients with the MFN2 variants outlined, stained for mtDNA (anti-dsDNA) in yellow, mitochondria (anti-TOMM20) in cyan and early endosomes (anti-RAB5C) in magenta. The top images are cropped in the yellow boxes and insets are shown accordingly. The yellow triangles indicate extra-mitochondrial mtDNA that colocalizes with Rab5 early endosomes, while the white circles show extra-mitochondrial mtDNA that is not in Rab5 early endosomes. (B) Quantification of average mitochondrial branch length in control versus R397_insertionKR patient fibroblasts. The violins show median mitochondrial branch length and IQR, n=3, each point indicates the total extra-mitochondrial mtDNA from 150 cells, colours indicate biological replicates. (C) Quantification of the relative mtDNA copy number in control versus R397_insertionKR patient fibroblasts. The bars show the mean +/- SD, n=3, with 8 technical replicates per biological replicate, each point indicates a replicate and colours indicate biological replicates. (D-J) Quantitative analyses of (D) numbers and (E) sizes of mtDNA nucleoids, number of extra-mitochondrial mtDNA puncta (F) co-localized with and (G) not co-localized with Rab5 early endosomes, (H) average distances between mitochondria and early endosomes and (I) numbers and (J) sizes of early endosomes per cell in U2OS WT/MFN2 KO and KO re-expression variants. For (D-J), the violin plots show the median and interquartile ranges, n=3, each replicate has 50 cells, the points show technical replicates and colours indicate the biological replicates. Statistics shown are one-way ANOVA, all statistics are compared to control fibroblasts, unless otherwise shown. Statistical symbols indicate- ns=P > 0.05, *=P ≤ 0.05, **= P ≤ 0.01, ****= P ≤ 0.0001.

## References

1. Al Khatib, I., Deng, J., Lei, Y., Torres-Odio, S., Rojas, G. R., Newman, L. E., Chung, B. K., Symes, A., Zhang, H., Huang, S.-Y. N., Pommier, Y., Khan, A., Shadel, G. S., West, A. P., Gibson, W. T., & Shutt, T. E. (2023). Activation of the cGAS-STING innate immune response in cells with deficient mitochondrial topoisomerase TOP1MT. Human Molecular Genetics, 32(15), 2422–2440. 10.1093/hmg/ddad062

2. Al Khatib, I., Deng, J., Symes, A., Kerr, M., Zhang, H., Huang, S.-Y. N., Pommier, Y., Khan, A., & Shutt, T. E. (2022). Functional characterization of two variants of mitochondrial topoisomerase TOP1MT that impact regulation of the mitochondrial genome. The Journal of Biological Chemistry, 298(10), 102420. 10.1016/j.jbc.2022.102420

3. Banchs, I., Casasnovas, C., Albertí, A., De Jorge, L., Povedano, M., Montero, J., Martínez-Matos, J. A., & Volpini, V. (2009). Diagnosis of Charcot-Marie-Tooth Disease. Journal of Biomedicine and Biotechnology, 2009, 985415. 10.1155/2009/985415

4. Baron, K. R., Oviedo, S., Krasny, S., Zaman, M., Aldakhlallah, R., Bora, P., Mathur, P., Pfeffer, G., Bollong, M. J., Shutt, T. E., Grotjahn, D. A., & Wiseman, R. L. (2025). Pharmacologic activation of integrated stress response kinases inhibits pathologic mitochondrial fragmentation. eLife, 13, RP100541. 10.7554/eLife.100541

5. Bora, P., Zaman, M., Oviedo, S., Kutseikin, S., Madrazo, N., Mathur, P., Pannikkat, M., Krasny, S., Aldakhlallah, R., Chu, A., Johnson, K. A., Grotjahn, D. A., Shutt, T. E., & Wiseman, R. L. (2025). Drug repurposing screen identifies an HRI activating compound that promotes adaptive mitochondrial remodeling in MFN2-deficient cells. Proceedings of the National Academy of Sciences of the United States of America, 122(48), e2517552122. 10.1073/pnas.2517552122

6. Boutant, M., Kulkarni, S. S., Joffraud, M., Ratajczak, J., Valera-Alberni, M., Combe, R., Zorzano, A., & Cantó, C. (2017). Mfn2 is critical for brown adipose tissue thermogenic function. The EMBO Journal, 36(11), 1543–1558. 10.15252/embj.201694914

7. Brožková, D. Š., Posádka, J., Laššuthová, P., Mazanec, R., Haberlová, J., Sišková, D., Sakmaryová, I., Neupauerová, J., & Seeman, P. (2013). Spectrum and frequencies of mutations in the MFN2 gene and its phenotypical expression in Czech hereditary motor and sensory neuropathy type II patients. Molecular Medicine Reports, 8(6), 1779–1784. 10.3892/mmr.2013.1730

8. Brunn, A., Zornbach, K., Hans, V. H., Haupt, W. F., & Deckert, M. (2012). Toll-like receptors promote inflammation in idiopathic inflammatory myopathies. Journal of Neuropathology and Experimental Neurology, 71(10), 855–867. 10.1097/NEN.0b013e31826bf7f3

9. Chen, H., Detmer, S. A., Ewald, A. J., Griffin, E. E., Fraser, S. E., & Chan, D. C. (2003). Mitofusins Mfn1 and Mfn2 coordinately regulate mitochondrial fusion and are essential for embryonic development. The Journal of Cell Biology, 160(2), 189–200. 10.1083/jcb.200211046

10. Choi, B.-O., Nakhro, K., Park, H. J., Hyun, Y. S., Lee, J. H., Kanwal, S., Jung, S.-C., & Chung, K. W. (2015). A cohort study of MFN2 mutations and phenotypic spectrums in Charcot-Marie-Tooth disease 2A patients. Clinical Genetics, 87(6), 594–598. 10.1111/cge.12432

11. Chung, K. W. (2006). Early onset severe and late-onset mild Charcot-Marie-Tooth disease with mitofusin 2 (MFN2) mutations. Brain, 129(8), 2103–2118. 10.1093/brain/awl174

12. Das, A., Nag, S., Mason, A. B., & Barroso, M. M. (2016). Endosome–mitochondria interactions are modulated by iron release from transferrin. The Journal of Cell Biology, 214(7), 831–845. 10.1083/jcb.201602069

13. David, J. O., Deng, J., Zaman, M., Swift, L., Shahhosseini, F., Sharma, A., Bureik, D., Padovani, F., Benedikt, A., Jaiswal, A., Mohan, A., Brideau, C., Grewal, S., Schmoller, K. M., Colarusso, P., & Shutt, T. E. (2026). A novel genetic fluorescent reporter to visualize mitochondrial nucleoids (p. 2023.10.23.563667). bioRxiv. 10.1101/2023.10.23.563667

14. de Brito, O. M., & Scorrano, L. (2008). Mitofusin 2 tethers endoplasmic reticulum to mitochondria. Nature, 456(7222), 605–610. 10.1038/nature07534

15. Dorn, G. W. (2020). Mitofusin 2 Dysfunction and Disease in Mice and Men. Frontiers in Physiology, 11, 782. 10.3389/fphys.2020.00782

16. Ershov, D., Phan, M.-S., Pylvänäinen, J. W., Rigaud, S. U., Le Blanc, L., Charles-Orszag, A., Conway, J. R. W., Laine, R. F., Roy, N. H., Bonazzi, D., Duménil, G., Jacquemet, G., & Tinevez, J.-Y. (2022). TrackMate 7: Integrating state-of-the-art segmentation algorithms into tracking pipelines. Nature Methods, 19(7), 829–832. 10.1038/s41592-022-01507-1

17. Estilow, T., Kozin, S. H., Glanzman, A. M., Burns, J., & Finkel, R. S. (2012). Flexor digitorum superficialis opposition tendon transfer improves hand function in children with Charcot-Marie-Tooth disease: Case series. Neuromuscular Disorders: NMD, 22(12), 1090–1095. 10.1016/j.nmd.2012.07.011

18. Fang, F., Marangoni, R. G., Zhou, X., Yang, Y., Ye, B., Shangguang, A., Qin, W., Wang, W., Bhattacharyya, S., Wei, J., Tourtellotte, W. G., & Varga, J. (2016). Toll-like Receptor 9 Signaling Is Augmented in Systemic Sclerosis and Elicits Transforming Growth Factor β-Dependent Fibroblast Activation. Arthritis & Rheumatology, 68(8), 1989–2002. 10.1002/art.39655

19. Franco, A., Li, J., Kelly, D. P., Hershberger, R. E., Marian, A. J., Lewis, R. M., Song, M., Dang, X., Schmidt, A. D., Mathyer, M. E., Edwards, J. R., Strong, C. de G., & Dorn, G. W. (2023). A human mitofusin 2 mutation can cause mitophagic cardiomyopathy. eLife, 12, e84235. 10.7554/eLife.84235

20. Gan, Q., Fu, X., Zhou, T., Fan, N., Nan, N., Wang, Y., Yang, Y., Gou, S., Hu, L., & Zhou, S. (2026). Mitochondrial DNA drives NLRP3-IL-1β axis activation in microglia by binding to NLRP3, leading to neurodegeneration in Parkinson’s disease models. Cell Death & Disease, 17(1), 213. 10.1038/s41419-026-08424-7

21. Gordaliza-Alaguero, I., Sànchez-Fernàndez-de-Landa, P., Radivojevikj, D., Villarreal, L., Arauz-Garofalo, G., Gay, M., Martinez-Vicente, M., Seco, J., Martín-Malpartida, P., Vilaseca, M., Macías, M. J., Palacin, M., Ivanova, S., & Zorzano, A. (2025). Endogenous interactomes of MFN1 and MFN2 provide novel insights into interorganelle communication and autophagy. Autophagy, 21(5), 957–978. 10.1080/15548627.2024.2440843

22. Hales, K. G., & Fuller, M. T. (1997). Developmentally regulated mitochondrial fusion mediated by a conserved, novel, predicted GTPase. Cell, 90(1), 121–129. 10.1016/s0092-8674(00)80319-0

23. Hammerling, B. C., Najor, R. H., Cortez, M. Q., Shires, S. E., Leon, L. J., Gonzalez, E. R., Boassa, D., Phan, S., Thor, A., Jimenez, R. E., Li, H., Kitsis, R. N., Dorn, G. W., Sadoshima, J., Ellisman, M. H., & Gustafsson, Å. B. (2017). A Rab5 endosomal pathway mediates Parkin-dependent mitochondrial clearance. Nature Communications, 8, 14050. 10.1038/ncomms14050

24. Hayashi, H., Saito, R., Tanaka, H., Hara, N., Koide, S., Yonemochi, Y., Ozawa, T., Hokari, M., Toyoshima, Y., Miyashita, A., Onodera, O., Okamoto, K., Ikeuchi, T., Nakajima, T., & Kakita, A. (2023). Clinicopathologic features of two unrelated autopsied patients with Charcot-Marie-Tooth disease carrying MFN2 gene mutation. Acta Neuropathologica Communications, 11(1), 207. 10.1186/s40478-023-01692-w

25. Hines, T. J., Bailey, J., Liu, H., Guntur, A. R., Seburn, K. L., Pratt, S. L., Funke, J. R., Tarantino, L. M., & Burgess, R. W. (2023). A Novel ENU-Induced Mfn2 Mutation Causes Motor Deficits in Mice without Causing Peripheral Neuropathy. Biology, 12(7), 953. 10.3390/biology12070953

26. Hori, M., Iwama, T., Asakura, Y., Kawanishi, M., Kamon, J., Hoshino, A., Takahashi, S., Takahashi, K., Nakaike, S., & Tsuruzoe, N. (2009). NT-702 (parogrelil hydrochloride, NM-702), a novel and potent phosphodiesterase 3 inhibitor, suppress the asthmatic response in guinea pigs, with both bronchodilating and anti-inflammatory effects. European Journal of Pharmacology, 618(1–3), 63–69. 10.1016/j.ejphar.2009.07.005

27. Irazoki, A., Gordaliza-Alaguero, I., Frank, E., Giakoumakis, N. N., Seco, J., Palacín, M., Gumà, A., Sylow, L., Sebastián, D., & Zorzano, A. (2023). Disruption of mitochondrial dynamics triggers muscle inflammation through interorganellar contacts and mitochondrial DNA mislocation. Nature Communications, 14(1), 108. 10.1038/s41467-022-35732-1

28. Jiang, J., Zuo, M., Zhao, K., Ling, Z., Wu, Z., Xue, D., Mo, S., Liu, Y., Chen, Y., Wang, J., Lu, B., Li, C., Duan, Y., He, H., & Song, Z. (2026). mtDNA leakage promotes neuron-glia crosstalk to induce epilepsy by cGAS-STING-driven neuroinflammation and serine metabolic reprogramming. Proceedings of the National Academy of Sciences of the United States of America, 123(9), e2522313123. 10.1073/pnas.2522313123

29. Lepelley, A., Della Mina, E., Van Nieuwenhove, E., Waumans, L., Fraitag, S., Rice, G. I., Dhir, A., Frémond, M.-L., Rodero, M. P., Seabra, L., Carter, E., Bodemer, C., Buhas, D., Callewaert, B., de Lonlay, P., De Somer, L., Dyment, D. A., Faes, F., Grove, L., … Crow, Y. J. (2021). Enhanced cGAS-STING-dependent interferon signaling associated with mutations in ATAD3A. The Journal of Experimental Medicine, 218(10), e20201560. 10.1084/jem.20201560

30. Liu, Y., Zhang, B., Duan, R., & Liu, Y. (2024). Mitochondrial DNA Leakage and cGas/STING Pathway in Microglia: Crosstalk Between Neuroinflammation and Neurodegeneration. Neuroscience, 548, 1–8. 10.1016/j.neuroscience.2024.04.009

31. McArthur, K., Whitehead, L. W., Heddleston, J. M., Li, L., Padman, B. S., Oorschot, V., Geoghegan, N. D., Chappaz, S., Davidson, S., San Chin, H., Lane, R. M., Dramicanin, M., Saunders, T. L., Sugiana, C., Lessene, R., Osellame, L. D., Chew, T.-L., Dewson, G., Lazarou, M., … Kile, B. T. (2018). BAK/BAX macropores facilitate mitochondrial herniation and mtDNA efflux during apoptosis. Science, 359(6378), eaao6047. 10.1126/science.aao6047

32. Montava-Garriga, L., Singh, F., Ball, G., & Ganley, I. G. (2020). Semi-automated quantitation of mitophagy in cells and tissues. Mechanisms of Ageing and Development, 185, 111196. 10.1016/j.mad.2019.111196

33. Newman, L. E., & Shadel, G. S. (2023). Mitochondrial DNA Release in Innate Immune Signaling. Annual Review of Biochemistry, 92, 299–332. 10.1146/annurev-biochem-032620-104401

34. Newman, L. E., Weiser Novak, S., Rojas, G. R., Tadepalle, N., Schiavon, C. R., Grotjahn, D. A., Towers, C. G., Tremblay, M.-È., Donnelly, M. P., Ghosh, S., Medina, M., Rocha, S., Rodriguez-Enriquez, R., Chevez, J. A., Lemersal, I., Manor, U., & Shadel, G. S. (2024). Mitochondrial DNA replication stress triggers a pro-inflammatory endosomal pathway of nucleoid disposal. Nature Cell Biology, 26(2), 194–206. 10.1038/s41556-023-01343-1

35. Nguyen, M., Collier, J. J., Ignatenko, O., Morin, G., Goyon, V., Janer, A., Tiefensee Ribeiro, C., Milnerwood, A. J., Huang, S., Desjardins, M., & McBride, H. M. (2025). MAPL regulates gasdermin-mediated release of mtDNA from lysosomes to drive pyroptotic cell death. Nature Cell Biology, 27(10), 1708–1724. 10.1038/s41556-025-01774-y

36. Oka, T., Hikoso, S., Yamaguchi, O., Taneike, M., Takeda, T., Tamai, T., Oyabu, J., Murakawa, T., Nakayama, H., Nishida, K., Akira, S., Yamamoto, A., Komuro, I., & Otsu, K. (2012). Mitochondrial DNA that escapes from autophagy causes inflammation and heart failure. Nature, 485(7397), 251–255. 10.1038/nature10992

37. Pipis, M., Feely, S. M. E., Polke, J. M., Skorupinska, M., Perez, L., Shy, R. R., Laura, M., Morrow, J. M., Moroni, I., Pisciotta, C., Taroni, F., Vujovic, D., Lloyd, T. E., Acsadi, G., Yum, S. W., Lewis, R. A., Finkel, R. S., Herrmann, D. N., Day, J. W., … Inherited Neuropathies Consortium - Rare Disease Clinical Research Network (INC-RDCRN). (2020). Natural history of Charcot-Marie-Tooth disease type 2A: A large international multicentre study. Brain: A Journal of Neurology, 143(12), 3589–3602. 10.1093/brain/awaa323

38. Ravindran, R., Velikkakath, A. K. G., Narendradev, N. D., Chandrasekharan, A., Santhoshkumar, T. R., & Srinivasula, S. M. (2022). Endosomal-associated RFFL facilitates mitochondrial clearance by enhancing PRKN/parkin recruitment to mitochondria. Autophagy, 18(12), 2851–2864. 10.1080/15548627.2022.2052460

39. Rodríguez-Nuevo, A., Díaz-Ramos, A., Noguera, E., Díaz-Sáez, F., Duran, X., Muñoz, J. P., Romero, M., Plana, N., Sebastián, D., Tezze, C., Romanello, V., Ribas, F., Seco, J., Planet, E., Doctrow, S. R., González, J., Borràs, M., Liesa, M., Palacín, M., … Zorzano, A. (2018). Mitochondrial DNA and TLR9 drive muscle inflammation upon Opa1 deficiency. The EMBO Journal, 37(10), e96553. 10.15252/embj.201796553

40. Rouzier, C., Bannwarth, S., Chaussenot, A., Chevrollier, A., Verschueren, A., Bonello-Palot, N., Fragaki, K., Cano, A., Pouget, J., Pellissier, J.-F., Procaccio, V., Chabrol, B., & Paquis-Flucklinger, V. (2012). The MFN2 gene is responsible for mitochondrial DNA instability and optic atrophy ‘plus’ phenotype. Brain: A Journal of Neurology, 135(Pt 1), 23–34. 10.1093/brain/awr323

41. Sawyer, S. L., Cheuk-Him Ng, A., Innes, A. M., Wagner, J. D., Dyment, D. A., Tetreault, M., Care4Rare Canada Consortium, Majewski, J., Boycott, K. M., Screaton, R. A., & Nicholson, G. (2015). Homozygous mutations in MFN2 cause multiple symmetric lipomatosis associated with neuropathy. Human Molecular Genetics, 24(18), 5109–5114. 10.1093/hmg/ddv229

42. Schindelin, J., Arganda-Carreras, I., Frise, E., Kaynig, V., Longair, M., Pietzsch, T., Preibisch, S., Rueden, C., Saalfeld, S., Schmid, B., Tinevez, J.-Y., White, D. J., Hartenstein, V., Eliceiri, K., Tomancak, P., & Cardona, A. (2012). Fiji: An open-source platform for biological-image analysis. Nature Methods, 9(7), 676–682. 10.1038/nmeth.2019

43. Sharma, G., Zaman, M., Sabouny, R., Joel, M., Martens, K., Martino, D., De Koning, A. P. J., Pfeffer, G., & Shutt, T. E. (2022). Characterization of a novel variant in the HR1 domain of MFN2 in a patient with ataxia, optic atrophy and sensorineural hearing loss. F1000Research, 10, 606. 10.12688/f1000research.53230.2

44. Stuppia, G., Rizzo, F., Riboldi, G., Del Bo, R., Nizzardo, M., Simone, C., Comi, G. P., Bresolin, N., & Corti, S. (2015). MFN2-related neuropathies: Clinical features, molecular pathogenesis and therapeutic perspectives. Journal of the Neurological Sciences, 356(1–2), 7–18. 10.1016/j.jns.2015.05.033

45. Tinevez, J.-Y., Perry, N., Schindelin, J., Hoopes, G. M., Reynolds, G. D., Laplantine, E., Bednarek, S. Y., Shorte, S. L., & Eliceiri, K. W. (2017). TrackMate: An open and extensible platform for single-particle tracking. Methods, 115, 80–90. 10.1016/j.ymeth.2016.09.016

46. Todkar, K., Chikhi, L., & Germain, M. (2019). Mitochondrial interaction with the endosomal compartment in endocytosis and mitochondrial transfer. Mitochondrion, 49, 284–288. 10.1016/j.mito.2019.05.003

47. Tripathi, A., Bartosh, A., Whitehead, C., & Pillai, A. (2023). Activation of cell-free mtDNA-TLR9 signaling mediates chronic stress-induced social behavior deficits. Molecular Psychiatry, 28(9), 3806–3815. 10.1038/s41380-023-02189-7

48. Uddin, G. M., Lacroix, R., Zaman, M., Khatib, I. A., Jaiswal, A., Rho, J. M., Kurrasch, D. M., & Shutt, T. E. (2026). The Ketogenic Diet Metabolite β-Hydroxybutyrate Promotes Mitochondrial Elongation via Deacetylation in HeLa Cells and Improves Autism-like Behavior in Zebrafish (p. 2022.10.03.510695). bioRxiv. 10.1101/2022.10.03.510695

49. Valente, A. J., Maddalena, L. A., Robb, E. L., Moradi, F., & Stuart, J. A. (2017). A simple ImageJ macro tool for analyzing mitochondrial network morphology in mammalian cell culture. Acta Histochemica, 119(3), 315–326. 10.1016/j.acthis.2017.03.001

50. VanPortfliet, J. J., Chute, C., Lei, Y., Shutt, T. E., & West, A. P. (2024). Mitochondrial DNA release and sensing in innate immune responses. Human Molecular Genetics, 33(R1), R80– R91. 10.1093/hmg/ddae031

51. Xian, H., Watari, K., Sanchez-Lopez, E., Offenberger, J., Onyuru, J., Sampath, H., Ying, W., Hoffman, H. M., Shadel, G. S., & Karin, M. (2022). Oxidized DNA fragments exit mitochondria via mPTP- and VDAC-dependent channels to activate NLRP3 inflammasome and interferon signaling. Immunity, 55(8), 1370–1385.e8. 10.1016/j.immuni.2022.06.007

52. Zaman, M., Chute, C., Marcadier, J. L., Grewal, S., Wiseman, R. L., Tyndall, A. V., McInnes, B., Innes, A. M., Bernier, F. P., & Shutt, T. E. (2026). A novel variant in MFN2 linked to a lethal disorder of neonatal onset (p. 2024.09.05.24313021). medRxiv. 10.1101/2024.09.05.24313021

53. Zaman, M., Sharma, G., Almutawa, W., Soule, T. G., Sabouny, R., Joel, M., Mohan, A., Chute, C., Joseph, J. T., Pfeffer, G., & Shutt, T. E. (2025). The MFN2 Q367H variant reveals a novel pathomechanism connected to mtDNA-mediated inflammation. Life Science Alliance, 8(6), e202402921. 10.26508/lsa.202402921

54. Zaman, M., & Shutt, T. E. (2022). The Role of Impaired Mitochondrial Dynamics in MFN2-Mediated Pathology. Frontiers in Cell and Developmental Biology, 10, 858286. 10.3389/fcell.2022.858286

55. Zecchini, V., Paupe, V., Herranz-Montoya, I., Janssen, J., Wortel, I. M. N., Morris, J. L., Ferguson, A., Chowdury, S. R., Segarra-Mondejar, M., Costa, A. S. H., Pereira, G. C., Tronci, L., Young, T., Nikitopoulou, E., Yang, M., Bihary, D., Caicci, F., Nagashima, S., Speed, A., … Frezza, C. (2023). Fumarate induces vesicular release of mtDNA to drive innate immunity. Nature, 615(7952), 499–506. 10.1038/s41586-023-05770-w

56. Zhang, H., He, Z., Yin, C., Yang, S., Li, J., Lin, H., Hu, G., Wu, A., Qin, D., Hu, G., & Yu, L. (2025). STING-mediated neuroinflammation: A therapeutic target in neurodegenerative diseases. Frontiers in Aging Neuroscience, 17, 1659216. 10.3389/fnagi.2025.1659216

