## Supplemental Figures for "Pathogenic Mitofusin 2 variants causing myopathy drive endosomal mtDNA release and inflammation"

### Supplemental Figure S1

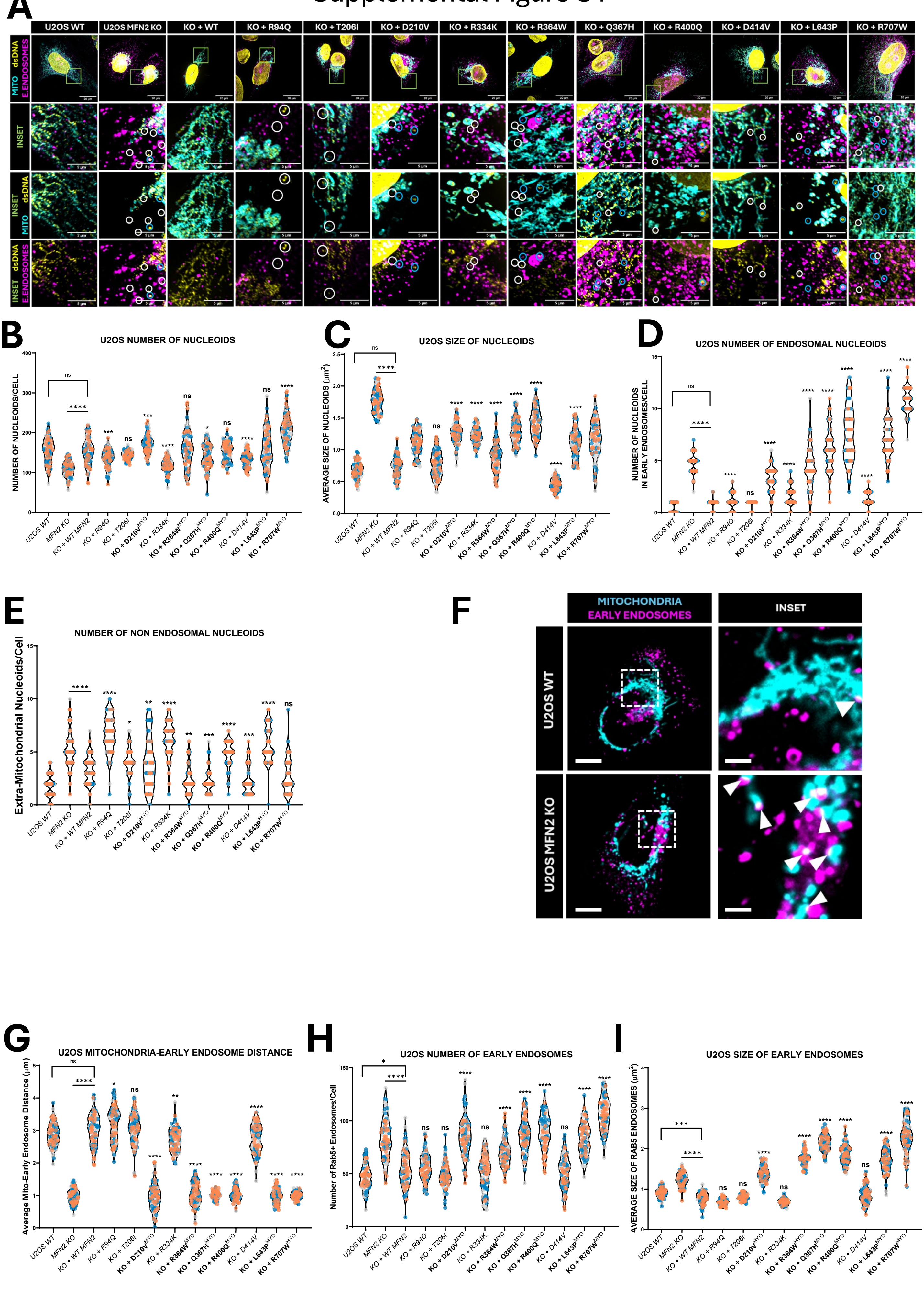

### Supplemental Figure 2

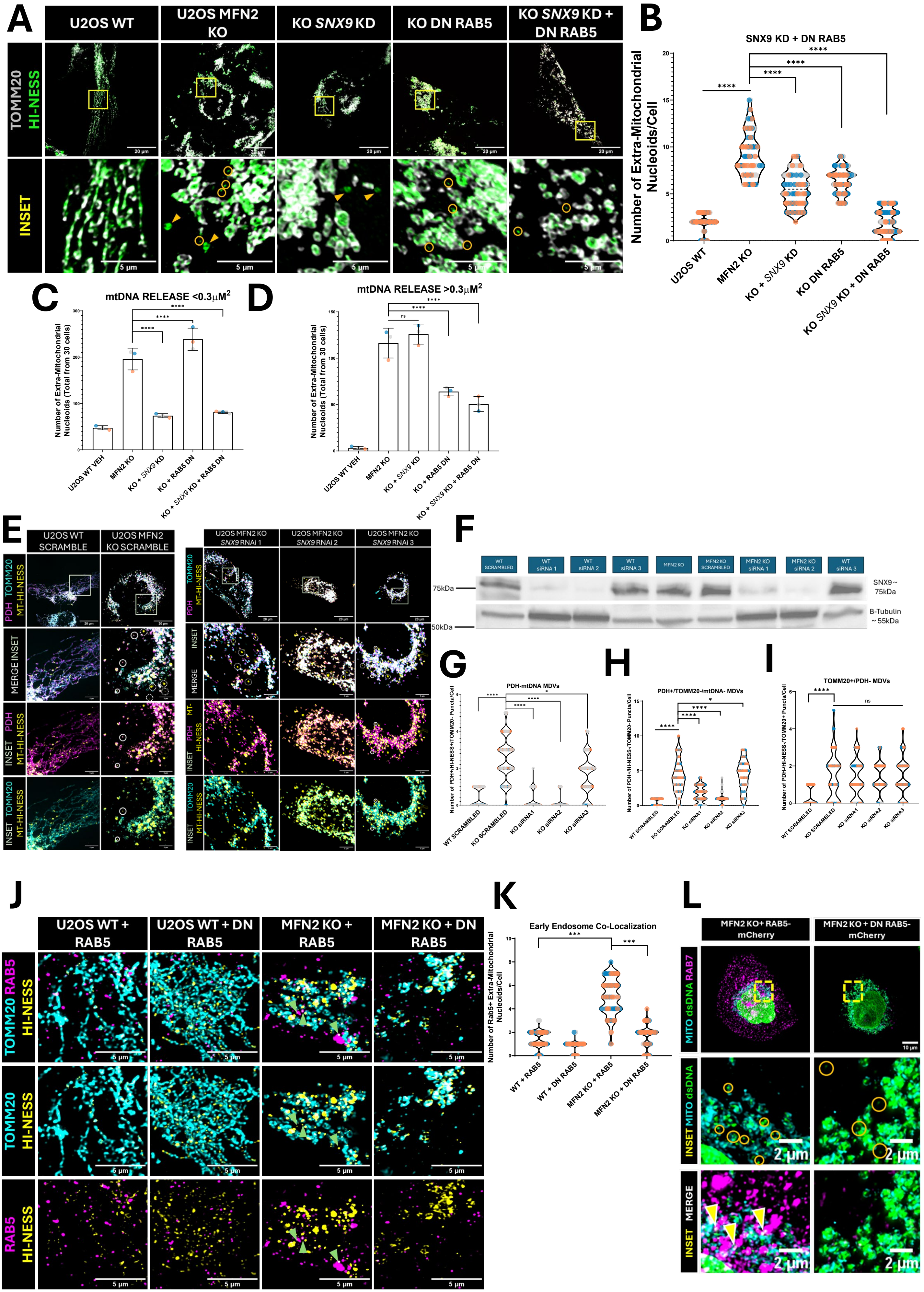

### Supplemental Figure S3

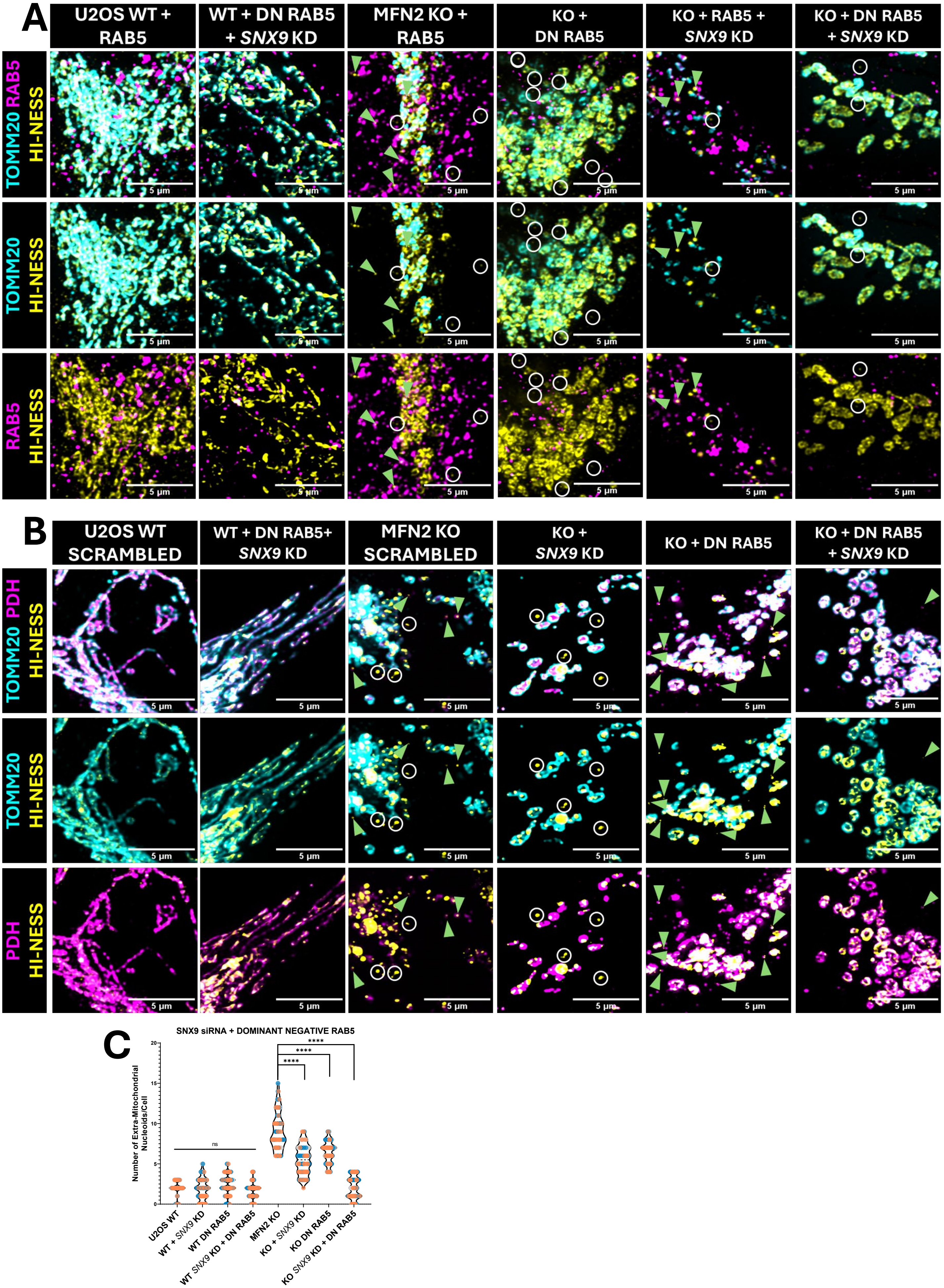

### Supplemental Figure S4

A

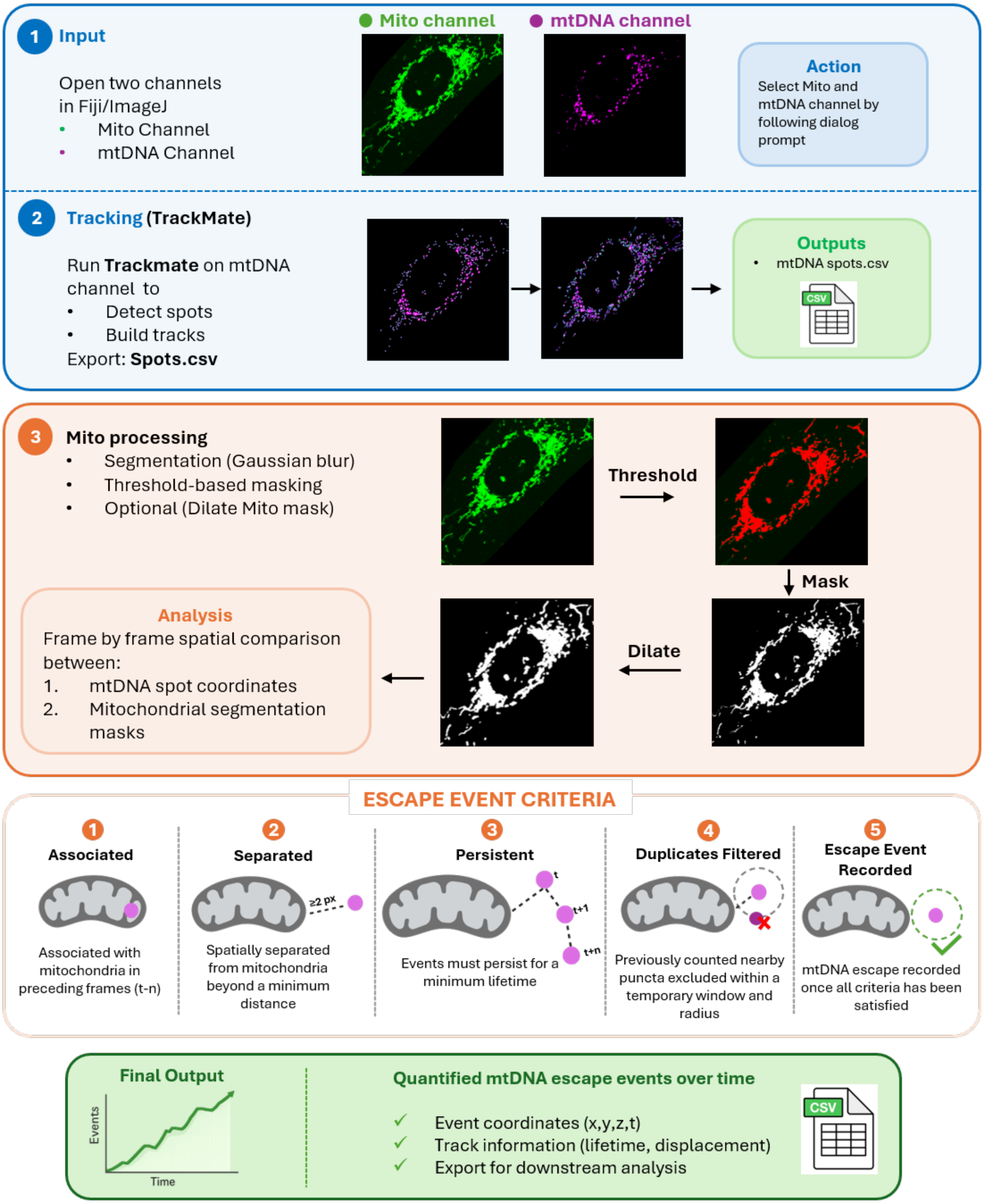

### Supplemental Figure S5

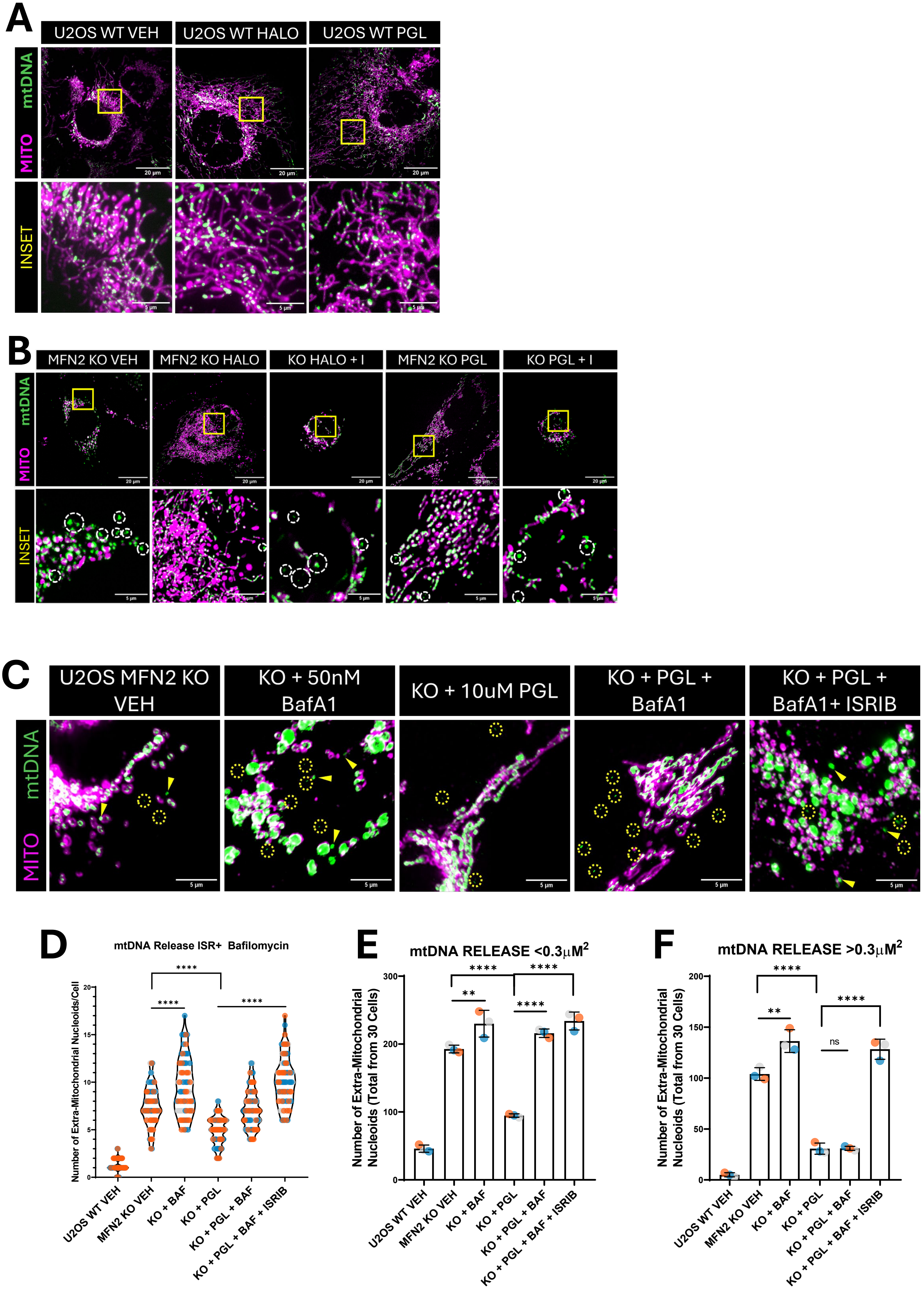

### Supplemental Figure S6

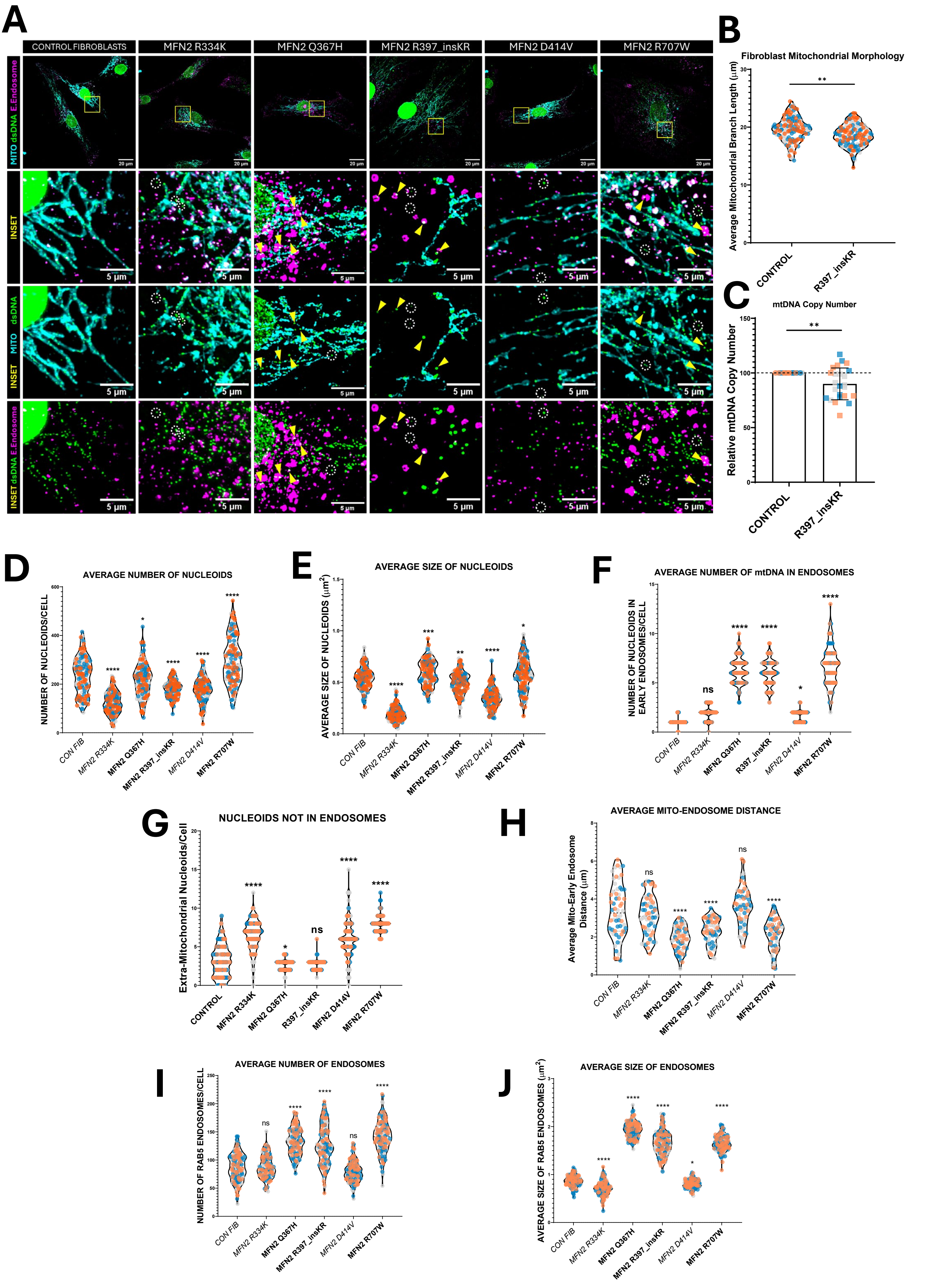
